# Antibody decay, T cell immunity and breakthrough infections following two SARS-CoV-2 vaccine doses in infliximab- and vedolizumab-treated patients

**DOI:** 10.1101/2021.11.10.21266168

**Authors:** Simeng Lin, Nicholas A Kennedy, Aamir Saifuddin, Diana Muñoz Sandoval, Catherine J Reynolds, Rocio Castro Seoane, Sherine H Kottoor, Franziska P Pieper, Kai-Min Lin, David K. Butler, Neil Chanchlani, Rachel Nice, Desmond Chee, Claire Bewshea, Malik Janjua, Timothy J McDonald, Shaji Sebastian, James L Alexander, Laura Constable, James C Lee, Charles D Murray, Ailsa L Hart, Peter M Irving, Gareth-Rhys Jones, Klaartje B Kok, Christopher A Lamb, Charlie W Lees, Daniel M Altmann, Rosemary J Boyton, James R Goodhand, Nick Powell, Tariq Ahmad, Contributors of the CLARITY IBD study

## Abstract

We report SARS-CoV-2 vaccine-induced immunity and risk of breakthrough infections in patients with inflammatory bowel disease treated with infliximab, a commonly used anti-TNF drug and those treated with vedolizumab, a gut-specific antibody targeting integrin a4b7 that does not impact systemic immunity. In infliximab-treated patients, the magnitude of anti-SARS-CoV2 antibodies was reduced 4-6-fold. One fifth of both infliximab- and vedolizumab-treated patients did not mount a T cell response. Antibody half-life was shorter in infliximab-treated patients. Breakthrough SARS-CoV-2 infections occurred more frequently in infliximab-treated patients and the risk was predicted by the level of antibody response after second vaccine dose. Overall, recipients of two doses of the BNT162b2 vaccine had higher anti-SARS-CoV-2 antibody concentrations, higher seroconversion rates, shorter antibody half-life and less breakthrough infections compared to ChAdOx1 nCoV-19 vaccine recipients. Irrespective of biologic treatment, higher, more sustained antibody levels were observed in patients with a history of SARS-CoV-2 infection prior to vaccination. Patients treated with anti-TNF therapy should be offered third vaccine doses.

## Introduction

Vaccination programmes have reduced SARS-CoV-2 transmission, hospitalisation and deaths^1^. Patients treated with immunosuppressive drugs were excluded from the original trials for COVID-19 vaccines^2,3^. Consequently, data relating to the magnitude and durability of immune responses and subsequent vaccine effectiveness in this population are limited^4^.

Drugs targeting tumor necrosis factor (TNF), such as infliximab, are the most frequently prescribed biological therapies used in the treatment of immune-mediated inflammatory disorders (IMIDs). Observational studies indicate that most patients with inflammatory bowel disease (IBD), an archetypal IMID, mount serological responses following SARS-CoV-2 vaccines, although most were underpowered to discern the impact of specific drugs, including immunomodulators (azathioprine, mercaptopurine, and methotrexate) and/or biologic therapies^5–8^. We reported that antibody responses following SARS-CoV-2 infection^9,10^ or a single-dose of either the BNT162b2 or ChAdOx1 nCoV-19 SARS-CoV-2 vaccines were impaired in anti-TNF treated patients when compared to vedolizumab treated patients^11^. Vedolizumab, is a gut-selective anti-integrin α4β7 monoclonal antibody that, unlike anti-TNF drugs is not associated with increased susceptibility to systemic infection or attenuated serological responses to vaccination^12^.

Here we compare immune responses between infliximab- and vedolizumab-treated patients with IBD who received two doses of the BNT162b2 or ChAdOx1 nCoV-19 vaccines. We hypothesised that, irrespective of the type of vaccine, antibody and T cell responses would be attenuated and less durable with an associated increased risk of breakthrough SARS-CoV-2 infections in patients treated with infliximab.

## Results

### Patient characteristics

Between September 22^nd^ 2020 and December 23^rd^ 2020, 7,226 patients were recruited to the CLARITY study from 92 UK hospitals^10^. In this analysis we included 2,264 infliximab- and 1,024 vedolizumab-treated participants without a history of prior SARS-CoV-2 infection, who had received uninterrupted biologic therapy since recruitment and had an antibody test between 14 and 70 days after a second-dose of the either the BNT162b2 and ChAdOx1 nCoV-19 SARS-CoV-2 vaccines. Participant characteristics are shown in Table 1.

**Table 1:** Baseline characteristics of participants who had anti-S RBD antibodies measured 2 to 10 weeks following 2 doses of COVID-19 vaccine. Abbreviations: IBD = inflammatory bowel disease; 5-ASA = 5-aminosalicylic acid; BMI = Body Mass Index; PRO2 = IBD disease activity. Values presented are median (interquartile range) or percentage (numerator/denominator). P values represent the results of a Mann Whitney U, Kruskal Wallis or Fisher’s exact test.

| Variable | Level | Vedolizumab | Infliximab | Overall | p |
| --- | --- | --- | --- | --- | --- |
| Vaccine | BNT162b2 | 40.0% (410/1024) | 40.1% (907/2264) | 40.1% (1317/3288) | 1.0 |
|  | ChAdOx1 nCoV-19 | 60.0% (614/1024) | 59.9% (1357/2264) | 59.9% (1971/3288) |  |
| Age (years) |  | 48.0 (35.1 - 61.5) | 40.2 (30.0 - 53.1) | 42.0 (31.3 - 55.7) | <0.0001 |
| Sex | Female | 48.0% (490/1021) | 45.9% (1038/2260) | 46.6% (1528/3281) | 0.16 |
|  | Male | 51.8% (529/1021) | 54.0% (1221/2260) | 53.3% (1750/3281) |  |
|  | Intersex | 0.0% (0/1021) | 0.0% (0/2260) | 0.0% (0/3281) |  |
|  | Prefer not to say | 0.2% (2/1021) | 0.0% (1/2260) | 0.1% (3/3281) |  |
| Ethnicity | White | 89.9% (917/1020) | 92.6% (2091/2259) | 91.7% (3008/3279) | 0.029 |
|  | Asian | 6.5% (66/1020) | 4.6% (103/2259) | 5.2% (169/3279) |  |
|  | Mixed | 2.5% (25/1020) | 1.4% (32/2259) | 1.7% (57/3279) |  |
|  | Black | 0.5% (5/1020) | 0.8% (18/2259) | 0.7% (23/3279) |  |
|  | Other | 0.7% (7/1020) | 0.7% (15/2259) | 0.7% (22/3279) |  |
| Diagnosis | Crohn's disease | 36.9% (378/1024) | 67.2% (1521/2264) | 57.8% (1899/3288) | <0.0001 |
|  | UC/IBDU | 63.1% (646/1024) | 32.8% (743/2264) | 42.2% (1389/3288) |  |
| Duration of IBD (years) |  | 9.0 (5.0 - 17.0) | 8.0 (3.0 - 16.0) | 8.5 (4.0 - 16.0) | 0.00015 |
| Age at IBD diagnosis (years) |  | 33.5 (22.8 - 47.2) | 27.6 (20.2 - 39.5) | 29.2 (20.8 - 42.1) | <0.0001 |
| Immunomodulators at vaccine |  | 20.5% (207/1009) | 57.7% (1295/2244) | 46.2% (1502/3253) | <0.0001 |
| 5-ASA |  | 33.8% (341/1009) | 20.6% (463/2244) | 24.7% (804/3253) | <0.0001 |
| Steroids |  | 5.9% (60/1009) | 2.8% (62/2244) | 3.8% (122/3253) | <0.0001 |
| BMI |  | 25.9 (23.0 - 29.8) | 25.9 (22.8 - 30.1) | 25.9 (22.9 - 30.0) | 0.81 |
| Heart disease |  | 4.8% (49/1020) | 2.6% (59/2256) | 3.3% (108/3276) | 0.0020 |
| Diabetes |  | 7.6% (78/1020) | 3.6% (82/2256) | 4.9% (160/3276) | <0.0001 |
| Lung disease |  | 16.5% (168/1020) | 13.1% (295/2256) | 14.1% (463/3276) | 0.011 |
| Kidney disease |  | 1.8% (18/1020) | 0.8% (17/2256) | 1.1% (35/3276) | 0.015 |
| Cancer |  | 1.6% (16/1020) | 0.3% (6/2256) | 0.7% (22/3276) | <0.0001 |
| Smoker | Yes | 8.1% (83/1020) | 10.8% (244/2256) | 10.0% (327/3276) | 0.00050 |
|  | Not currently | 37.6% (384/1020) | 30.2% (682/2256) | 32.5% (1066/3276) |  |
|  | Never | 54.2% (553/1020) | 59.0% (1330/2256) | 57.5% (1883/3276) |  |
| Exposure to documented cases of COVID-19 |  | 7.8% (80/1020) | 8.6% (194/2256) | 8.4% (274/3276) | 0.50 |
| Income deprivation score |  | 0.089 (0.055 - 0.146) | 0.091 (0.052 - 0.153) | 0.090 (0.053 - 0.151) | 0.86 |
| Active disease (PRO2) |  | 10.5% (100/956) | 5.1% (109/2132) | 6.8% (209/3088) | <0.0001 |
| Time between vaccine doses (weeks) |  | 10.9 (9.7 - 11.1) | 11.0 (10.0 - 11.3) | 10.9 (10.0 - 11.3) | 0.0042 |
| Time from second dose to serum sample (weeks) |  | 5.7 (3.7 - 7.7) | 5.7 (3.7 - 7.7) | 5.7 (3.7 - 7.7) | 0.74 |

Additional analyses are presented for a subset of 211 infliximab- and 71 vedolizumab-treated patients included in our T cell experiments (Supplementary Table 2), and a further 525 infliximab- and 224 vedolizumab-treated participants who had a history of SARS-CoV-2 infection before vaccination (Supplementary Table 3).

### Anti-SARS-CoV-2 (S) antibody level following second COVID-19 vaccine

Overall, the geometric mean [geometric SD] of anti-S RBD antibody concentration was higher in recipients of two doses of the BNT162b2 than ChAdOx1 nCoV-19 vaccines (1080.2 U/mL [7.6] vs 289.9 U/mL[5.2], p < 0.0001). Anti-S RBD antibody concentrations were lower in patients treated with infliximab than in those treated with vedolizumab, following a second dose of BNT162b2 (565.1 U/mL [6.2] vs 4527.6 U/mL [5.4], p <0.0001) and ChAdOx1 nCoV-19 (184.5 U/mL [5.0] vs 786.2 U/mL [3.5], p <0.0001) vaccines (Fig. 1).

**Figure 1:**
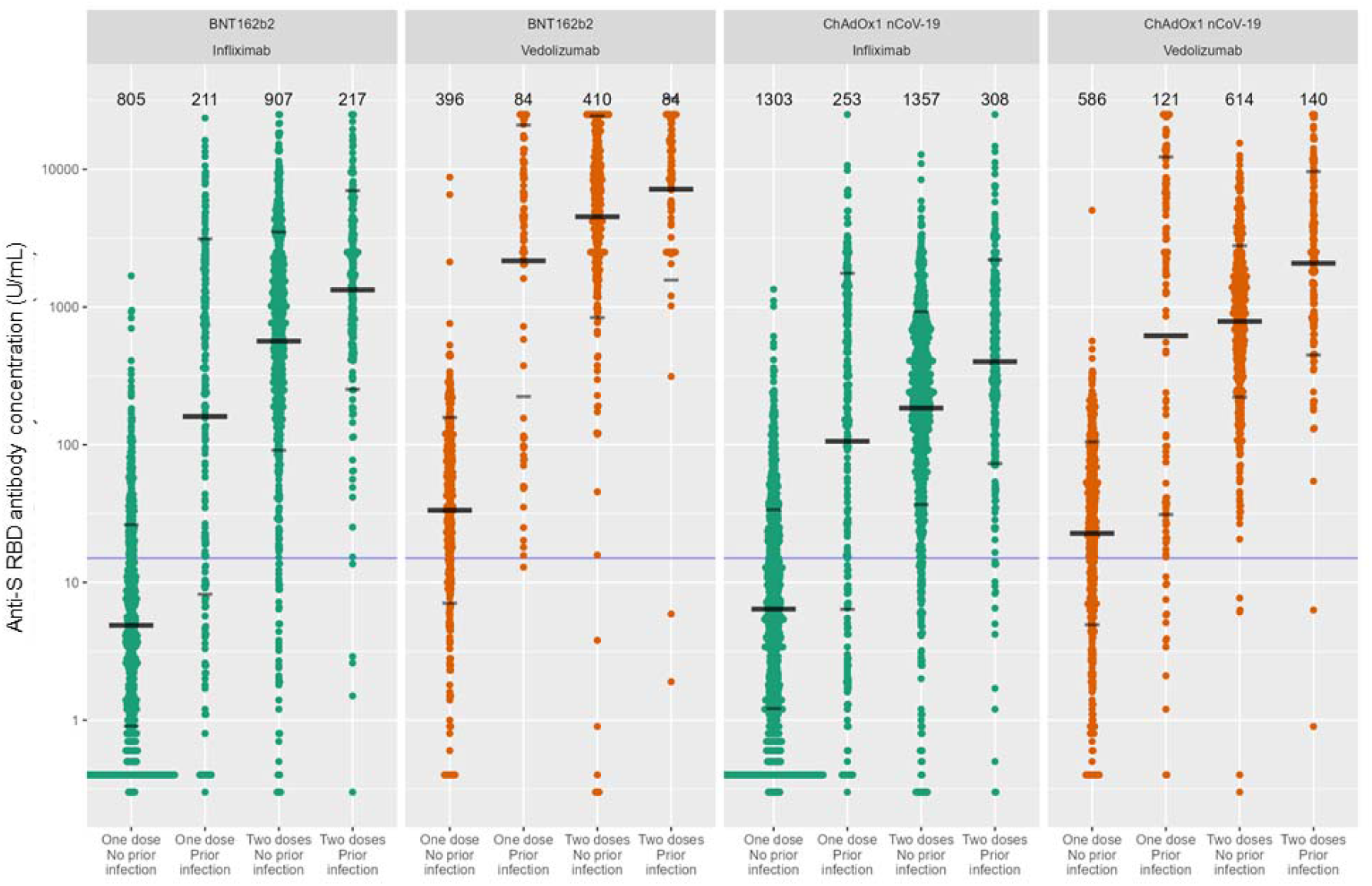
Anti-S RBD antibody concentration stratified by biologic therapy (infliximab vs vedolizumab), type of vaccine, vaccine dose and history of prior SARS-CoV-2 infection. The wider bar represents the geometric mean, while the narrower bars are drawn one geometric standard deviation either side of the geometric mean. Based on published data using neutralization assays threshold shown of 15 U/mL was used to determine seroconversion^11^. The biologic treatment infliximab is shown in green and vedolizumab in orange. The number of individuals tested for each group are shown in black at the top of each panel.

Crude sensitivity analyses, excluding patients treated with a concomitant immunomodulator, confirmed lower anti-S RBD antibody concentrations in patients treated with infliximab alone versus vedolizumab alone (BNT162b2 816.5 U/mL [4.9] vs 4616.3U/mL [6.0], p <0.0001, ChAdOx1 nCoV-19 181.4 U/mL [4.4] vs 774.9 U/mL [3.6], p <0.0001).

After propensity matching for immunomodulator use and the other factors associated with choice of biologic, we confirmed lower anti-S RBD antibody concentrations in infliximab-compared to vedolizumab-treated patients (BNT162b2 587.9 U/mL [6.1] vs 4657.5U/mL [4.7], p <0.0001, ChAdOx1 nCoV-19 191.1 U/mL [4.7] vs 778.6 U/mL [3.7], p <0.0001) (Supplementary Table 4).

Multivariable linear regression analyses in patients without prior SARS-CoV-2 infection confirmed that antibody concentrations were reduced four and six-fold in infliximab-compared with vedolizumab-treated participants who received the BNT162b2 (fold change [FC] 0.15 [95% CI 0.12, 0.19], p<0.0001) and ChAdOx1 nCoV-19 ([FC] 0.24 [95% CI 0.20, 0.28], p<0.0001) vaccines (Fig. 2a and Fig. 2b respectively). Age ≥60 years and Crohn’s disease were also independently associated with lower anti-S RBD antibody concentrations in vaccinated participants. Thiopurine or methotrexate use was independently associated with lower anti-S RBD antibody concentrations in participants who received the BNT162b2, but not the ChAdOx1 nCoV-19 vaccine. Current smoking, non-white ethnicity and steroid use were associated with lower anti-S RBD antibody concentrations in participants who received the ChAdOx1 nCoV-19 but not the BNT162b2 vaccine. To assess the effect of vaccine type on antibody responses, we combined our response data in a model that included vaccine type in addition to the significant factors above. Vaccination with the BNT162b2 vaccine compared to the ChAdOx1 nCoV-19 was independently associated with a 3.7 fold [95% CI 3.28 – 4.12] higher anti-S RBD antibody concentration (p < 0.0001) (Fig. 2c).

**Figure 2:**
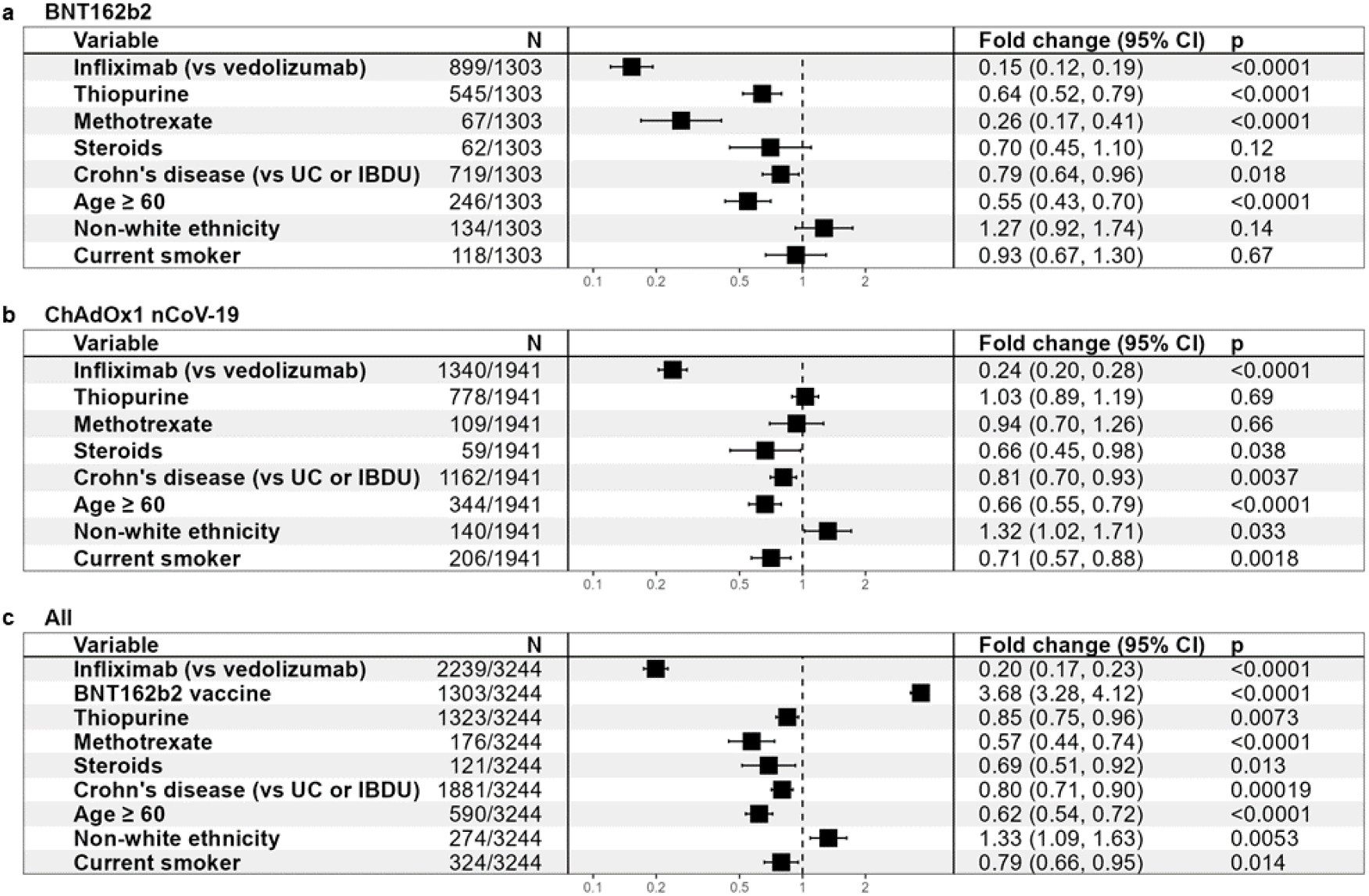
Exponentiated coefficients of linear regression models of log anti-S RBD antibody concentration. Exponentiated coefficients of linear regression model of log anti-S RBD antibody concentration in participants who received **a**. BNT162b2 vaccine. **b**. ChAdOx1 nCoV-19 vaccine. c. either the BNT162b2 or ChAdOx1 nCoV-19 vaccine. The resultant values represent the fold change of antibody concentration associated with each variable. Each vaccine was modelled separately, and then a further model was created using all available data. Horizontal dotted line represents a fold change of 1. Abbreviations: UC = ulcerative colitis, IBDU = IBD unclassified

Seroconversion rates after the first vaccine dose were lower in infliximab-compared to vedolizumab-treated participants (Fig. 1). However, administration of a second vaccine dose resulted in a >100-fold and >25 fold increase in antibody concentrations in recipients of the BNT162b2 and ChAdOx1 nCoV-19 vaccines, respectively (Fig. 1). Overall, more infliximab-than vedolizumab-treated patients failed to seroconvert after their second vaccine dose (5.9% vs 1.3%, p < 0.0001). Seroconversion rates stratified by biologic therapy and vaccine type are reported in Supplementary Fig 1.

### Anti-spike T cell responses following two doses of BNT162b2 and ChAdOx1 nCoV-19 SARS-CoV-2 vaccines

There were no significant differences in the magnitude of anti-spike T cell responses observed in infliximab-compared with vedolizumab-treated patients after one or two doses of either vaccine (Fig. 3a). The proportion of patients failing to mount detectable T cell responses were similar in both groups (infliximab 19.6% vs. vedolizumab 19.2%). For recipients of one and two doses of BNT162b2 vaccine there was a modest positive correlation between T cell responses and antibody concentration. This association was not observed in recipients following either dose of the ChAdOx1 nCoV-19 vaccine (Fig. 3b). When T cell responses were ranked by magnitude of antibody responses, most patients who did not mount an antibody response after the first vaccine dose (indicated by the dark grey bar) had a detectable T cell response (Fig. 4). In addition to the uncoupling of the T cell and antibody responses demonstrated, this analysis emphasised that about one fifth of participants made no T cell responses irrespective of vaccine used (indicated by the light grey bars). Moreover, a minority of individuals (3/67) 4.5% for BNT162b2 and (1/56) 1.8% for ChAdOx1 nCoV-19 vaccines carry neither detectable antibody nor T cell responses after two doses of vaccine (Fig. 3b, Fig. 4).

**Figure 3.**
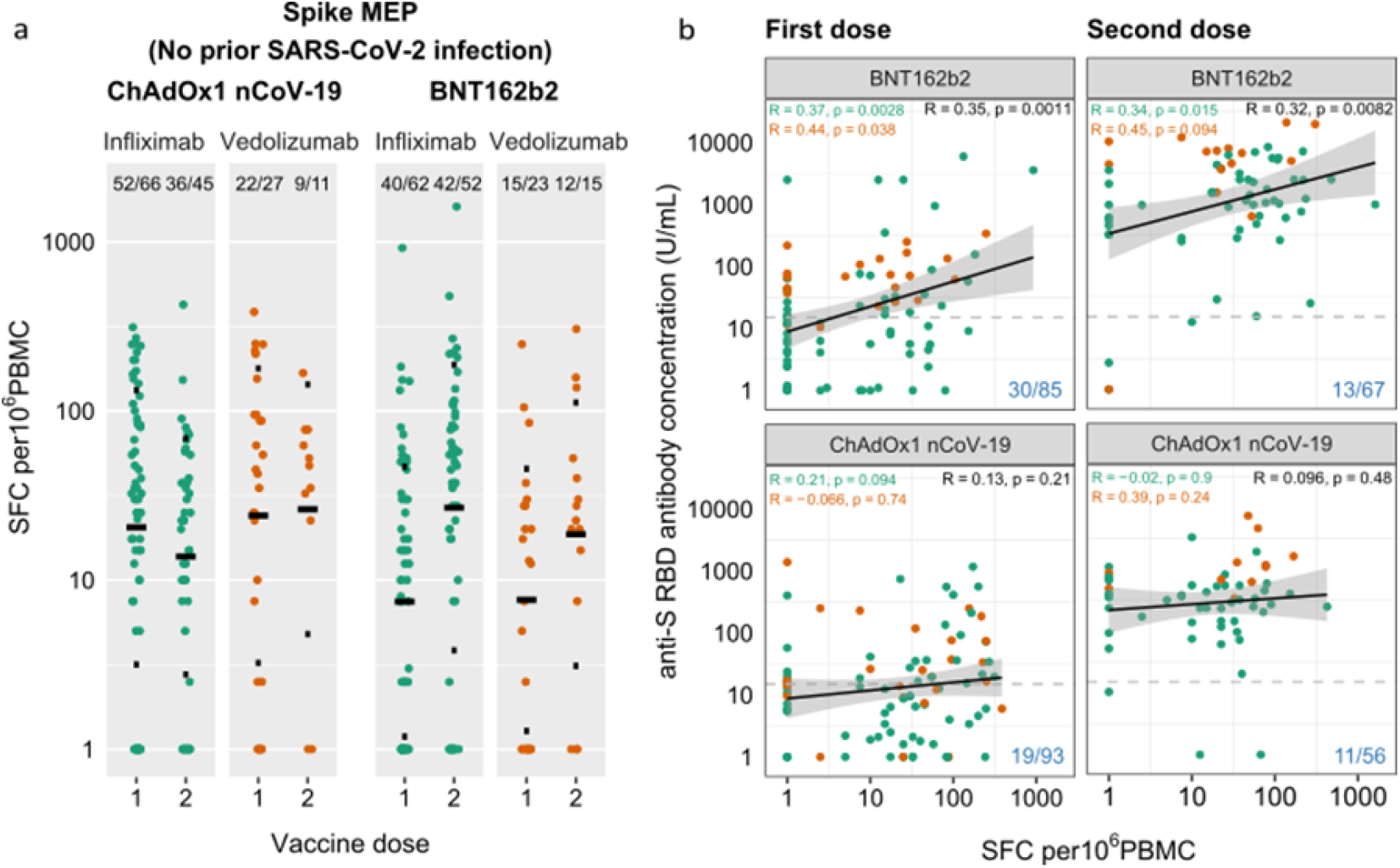
Anti-SARS-CoV-2 spike T cell responses stratified by vaccine platform (BNT162b2 vs ChAdOx1 nCoV-19), biologic therapy (infliximab vs vedolizumab), and vaccine dose (one vs two). **a**. Spike MEP T cell responses SFC per 10^6^ PBMC stratified by vaccine platform, biologic therapy (infliximab vs vedolizumab) and number of vaccine doses. The horizontal bar represents the geometric mean and the narrow bars represent one geometric standard deviation either side of the geometric mean. The number of T cell responders / total number of individuals tested are shown in black at the top of each panel. **b**. Scatterplot demonstrating the correlation between T cell responses against spike MEP pool (SFC per 10^6^ PBMC) and anti-SARS-CoV-2 spike antibody concentration after the first (LHS) and second (RHS) dose of BNT162B2 (top) and ChAdOx1 nCoV-19 (bottom) vaccine. The number of non-T cell responders / total number of individuals tested is shown in blue on the bottom RHS of each panel. The horizontal dotted line in **b**. represents a threshold of 15 U/mL of anti-S1 SARS-CoV-2 antibody. The biologic infliximab is show in green and vedolizumab is shown in orange. R, Spearman’s rank correlation. SFC, spot forming cells. PBMC, peripheral blood mononuclear cell. MEP, mapped epitope peptide.

**Figure 4:**
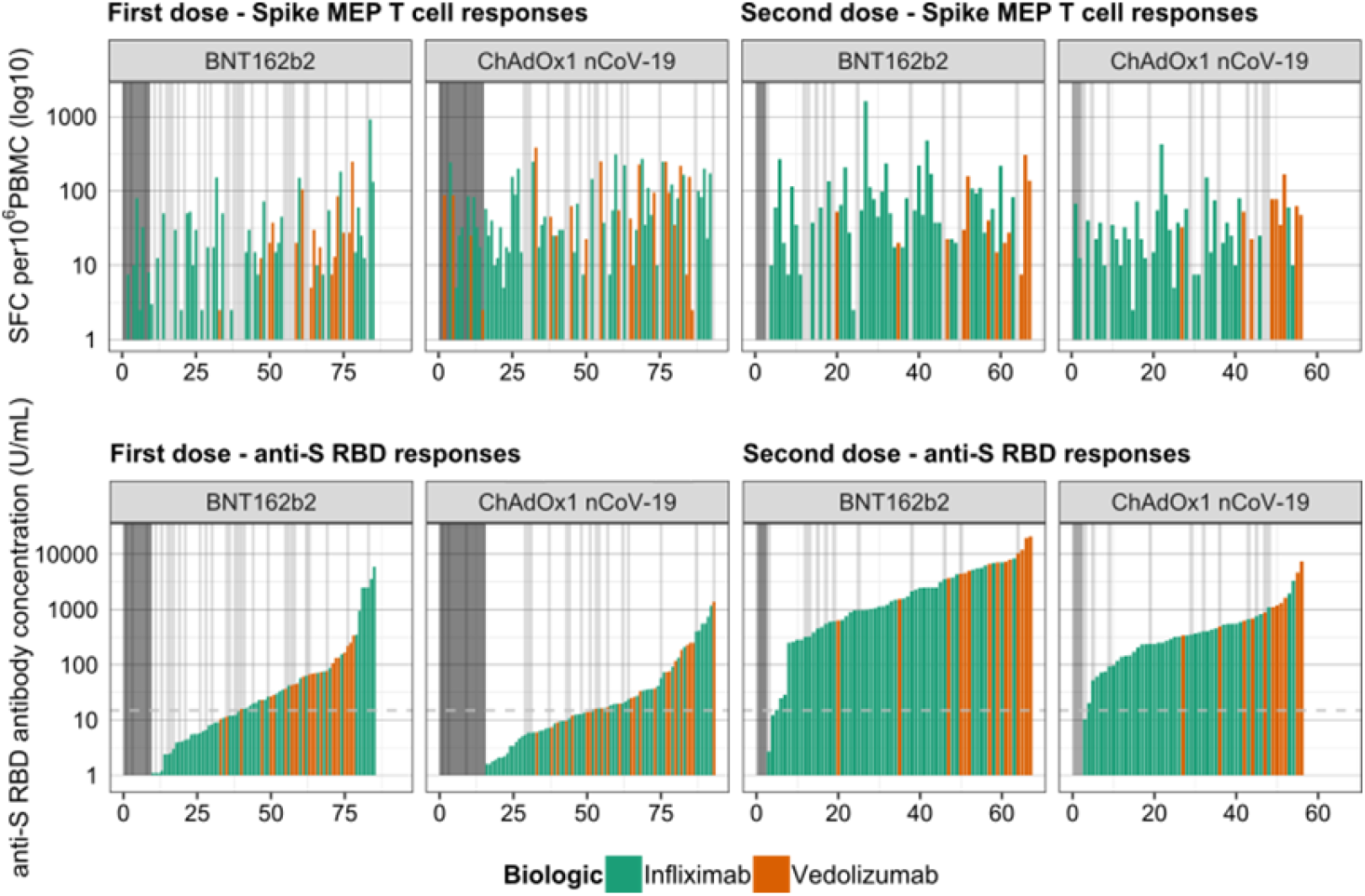
Anti-spike T cell responses ordered by cumulative magnitude of anti-S RBD following two doses of the BNT162b2 or ChAdOx1 nCoV-19 vaccine shows uncoupling of the T cell and antibody responses. Top panel shows T cell responses to spike, and bottom panel shows anti-S RBD responses plotted for individual study participants ordered by increasing magnitude of anti-S RBD antibody concentration (U/mL). The vertical dark grey bars at the LHS of the panels indicate individuals with no anti-S RBD response. The vertical light grey bars in the panels indicate individuals with no T cell response. The horizontal dotted line represents a threshold shown of 15 U/mL of anti-S RBD.

### Durability of antibody responses following two doses of BNT162b2 and ChAdOx1 nCoV-19 SARS-CoV-2 vaccines

The estimated half-life of anti-S RBD antibodies was shorter in participants receiving the BNT162b2 compared to the ChAdOx1 nCoV-19 vaccines (4.6 weeks [95% CI 4.44 – 4.74] vs 5.9 weeks [95% CI 5.88 – 6.29], p value < 0.0001). When stratified by biologic, half-life estimates were shorter in infliximab-than vedolizumab-treated patients following two-doses of BNT162b2 (4.0 weeks [95% CI 3.8 – 4.1] vs 7.2 weeks [95% CI 6.8 – 7.6]) and ChAdOx1 nCoV-19 (5.3 weeks [95% CI 5.1 – 5.5] vs 9.3 weeks [95% CI 8.5 – 10.2], p value < 0.0001) (Supplementary Fig. 2 and Supplementary Table 5). Overall, following two doses of either vaccine, anti-S RBD antibodies showed minimal decay to last follow-up in patients treated with vedolizumab (Fig. 5) and were similar to those observed in participants in the Virus Watch community cohort (Supplementary Fig. 4). However, in infliximab-treated participants the geometric mean concentrations dropped to the seroconversion threshold by about 25 weeks after the second dose irrespective of vaccine administered (Fig. 5). Infliximab compared to vedolizumab treatment, current smoking and white ethnicity were associated with a faster fall in anti-S RBD antibody concentration below the seroconversion threshold. (Supplementary Fig. 5, 6).

**Figure 5:**
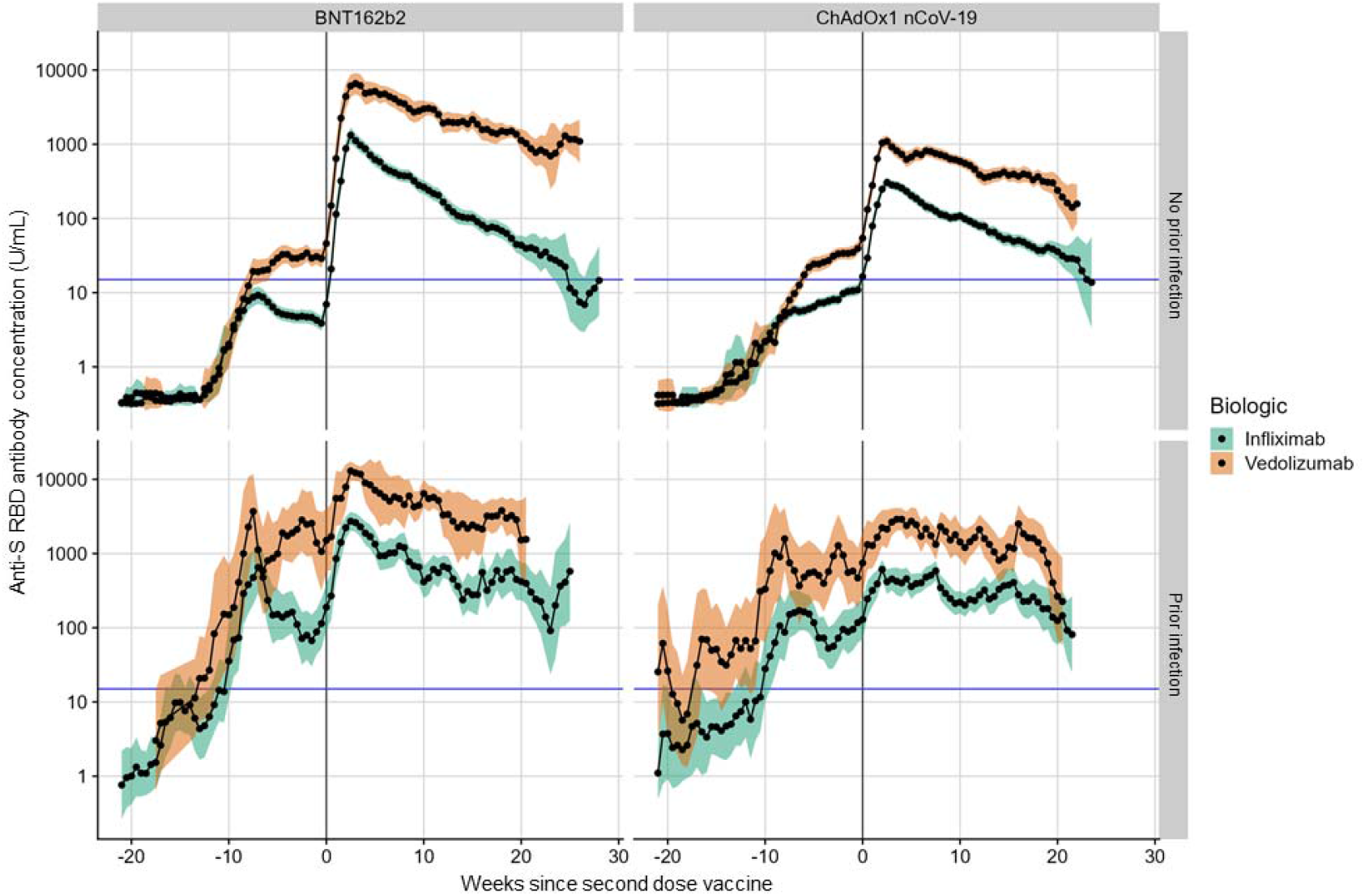
Rolling geometric mean antibody concentration over time from the date of the second dose of the SARS-CoV-2 vaccine (week 0) stratified by biologic therapy (infliximab vs vedolizumab), vaccine, and history of prior SARS-CoV-2 infection. Geometric means are calculated using a rolling 15-day window (i.e. 7 days either side of the day indicated). The shaded areas represent the 95% confidence intervals of the geometric means. The horizontal blue line represents the seroconversion threshold (15 U/mL). The number of participants included at each time point is presented in Supplementary Figure 3. Overall, data from 4429 participants with no history of prior infection (2999 on infliximab and 1430 on vedolizumab) and 1170 participants with a history of prior infection (825 on infliximab and 345 on vedolizumab) were included in this graph between 22 weeks before and 29 weeks after the second vaccine dose. The biologic treatment infliximab is shown in green and vedolizumab is shown in orange.

**Figure 6:**
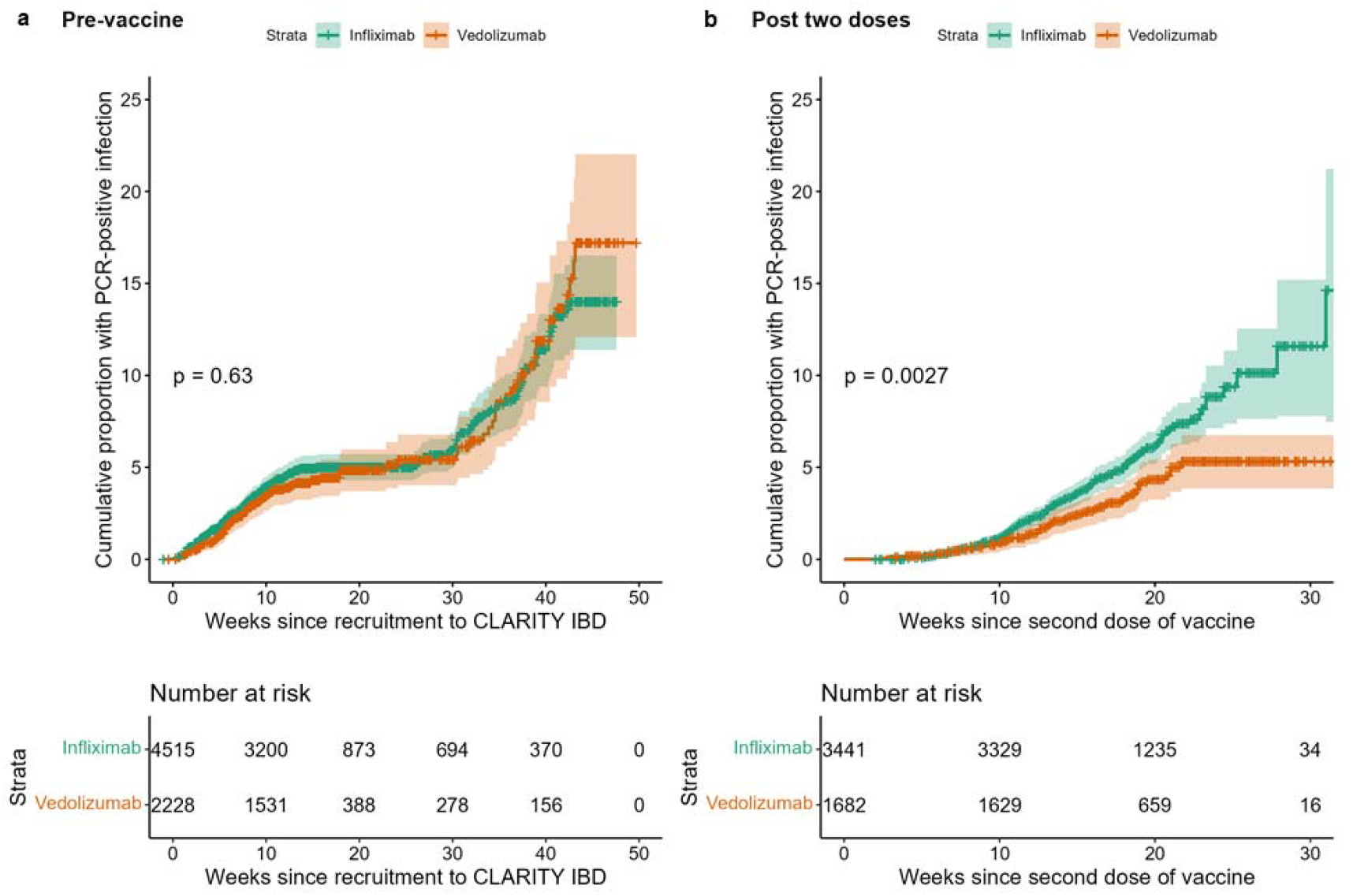
Kaplan-Meier graphs comparing the time to PCR-confirmed SARS-CoV-2 infection stratified by biologic therapy (infliximab vs vedolizumab) in participants before vaccination and after receiving two doses of vaccine. **a)** The time to PCR-confirmed SARS-CoV-2 infection in participants who have not received any dose of either vaccine stratified by biologic therapy (infliximab vs vedolizumab). **b)** The time to a PCR-confirmed SARS-CoV-2 breakthrough infection in participants following two doses of either vaccine stratified by biologic therapy. The biologic treatment infliximab is shown in green and vedolizumab in orange. The number of participants at each time point are displayed in black at the bottom of each figure. P-values are calculated using log-rank test.

### Breakthrough SARS-CoV-2 infections following two doses of vaccine

267/5123 participants without PCR-positive or serological evidence of prior SARS-CoV-2 infection had a first positive SARS-CoV-2 PCR test two or more weeks after the second vaccine dose. Overall, 89.2% patients were symptomatic: the most commonly reported symptoms were fatigue (73.7%), anosmia/ageusia (71.4%), fever (57.1%), cough (54.9%), myalgia (45.9%), hoarse voice (30.8%), confusion (27.8%) and chest pains (23.3%). 1.2% (3/251) of participants with PCR confirmed infection were hospitalised because of COVID-19.

Breakthrough SARS-CoV-2 infections were more frequent (5.8% (201/3441) vs 3.9% (66/1682), p = 0.0039) and the time to breakthrough shorter in patients treated with infliximab than vedolizumab (p = 0.0027) (Fig. 6b). In contrast biologic class did not impact on time to PCR confirmed infection prior to vaccination (p = 0.63) (Fig. 6a). In a model that included biologic and vaccine type, shorter time to breakthrough infection was associated with infliximab (Hazard Ratio (HR) 1.52 [95% CI 1.15 – 2.01], p = 0.003) and having received the ChAdOx1 nCoV-19 (HR 1.49 [95% CI 1.15 – 1.92], p = 0.0023) vaccine. Geometric mean [geometric SD] anti-S RBD antibody concentrations measured 2 to 10 weeks after a second vaccine dose were significantly lower in participants who subsequently had a PCR confirmed breakthrough SARS-CoV-2 infection: for every 10-fold rise in anti-S RBD antibody level we observed a 0.8-fold reduction in odds of breakthrough infection ([95% CI 0.70 – 0.99], p = 0.03).

### Antibody responses in patients with prior SARS-CoV-2 infection

Amongst patients with a history of SARS-CoV-2 infection before vaccination, geometric mean [SD] anti-S RBD antibody concentrations were lower in infliximab-compared with vedolizumab-treated patients after a second dose of BNT162b2 (1330.0 U/mL [5.3] vs 7169.5 U/mL [4.6], p <0.0001) and ChAdOx1 nCoV-19 (401.2 U/mL [5.5] vs 2077.3 [4.6] p <0.0001) vaccines. In all patients, antibody concentrations following vaccination were higher in patients without a history of SARS-CoV-2 infection (Fig. 1). Irrespective of vaccine or biologic type, minimal decay of anti-S RBD antibodies were observed up to a follow-up of 21 weeks.

## Discussion

We have shown that in infliximab-treated patients, anti-SARS-CoV-2 spike antibody responses are attenuated following two doses of the BNT162b2 and ChAdOx1 nCoV-19 SARS-CoV-2 vaccines. One fifth of both infliximab- and vedolizumab-treated patients did not mount a T cell response and a small subset of patients had neither antibody nor T cell responses. Antibody half-lives were shorter in infliximab treated patients. Breakthrough SARS-CoV-2 infections were more common and occurred earlier in infliximab-treated patients who received the ChAdOx1 nCoV-19 vaccine. The risk of breakthrough infection was predicted by lower antibody levels after the second dose of vaccine. Irrespective of biologic treatment, higher and more sustained antibody levels, were observed in patients with a history of SARS-CoV-2 infection.

Sustained antibody responses observed in vaccinated patients with a history of prior SARS-CoV-2 infection indicates that third antigen exposure enhances the serological response. This supports the rationale for prioritising a third dose of vaccine to clinically vulnerable patient populations^13–16^, who otherwise may face further periods of social distancing or hospitalisation following infection. Whilst drawing direct comparisons between IBD patients and patients treated with more potent chemotherapies is limited by the degree to which patients are immunosuppressed, data from solid organ transplant recipients shows that a third dose of vaccine also leads to sustained immune responses^17^.

Irrespective of biologic or immunosuppressant use, and in keeping with the original trials^2,18^, the highest antibody responses were seen in recipients of the BNT162b2 vaccine. Like in the general population these responses waned more quickly than in the recipients of the ChAdOx1 nCoV-19 vaccine^19^. Unlike the general population^20^, but similar to renal transplant recipients^4^, we did not observe differences in T cell ELISpot responses between recipients of the BNT162b2 and ChAdOx1 nCoV-19 vaccines. The differences observed in breakthrough infection by vaccine type reported here are consistent with the differences in efficacy reported in the respective clinical trials^2,3,21^. The higher peak antibody levels and the lower rate of SARS-CoV-2 breakthrough infections suggest that the BNT162b2 rather than the ChAdOx1 nCoV-19 vaccine should be used for primary vaccination in infliximab-treated patients and although untested supports the use of BNT162b2 for third doses in all patients treated with an anti-TNF regardless of the primary vaccine type.

All patients treated with anti-TNF therapy should receive a third primary dose of SARS-CoV-2 vaccine and our data support recent recommendations that this should occur about 4-8 weeks after the second dose^13,14,16^ during periods of high transmission in the population. Our data demonstrate that patients treated with vedolizumab and infliximab-treated patients with prior SARS-CoV-2 infection have sustained antibody levels beyond 6 months.

When starting a biologic, it would be reasonable to consider differences in SARS-CoV-2 vaccine response as one of the factors when determining which drug to use. For patients who need to start anti-TNF therapy, the benefits of combination immunomodulator therapy should be weighed against the risk of attenuated vaccine response and whenever feasible, patients should first receive a SARS-CoV-2 vaccine dose. Further research to determine whether timing third vaccine doses towards the end of anti-TNF treatment cycles when drug levels are lowest leads to greater immunogenicity^9^ is needed. Other strategies including the temporary discontinuation of immunomodulators^22^, the use of heterologous vaccines^23^ and adjuvants including the influenza vaccines (ComFluCOV)^24^ need to be studied in immunosuppressed patient groups.

The biology underpinning loss of durable antibody responses and uncoupling of the B cell and T cell responses merit further research. TNF is a pleiotropic cytokine and its activities include maturation of antigen presenting cells, modulation of T cell responses and stimulation of immunoglobulin synthesis^25–27^. TNF neutralization, or genetic ablation, results in substantial loss of B-cells in primary follicles in germinal centres, reduced numbers of memory B-cells in the periphery but preserved numbers of T cells^25^. Uncoupling of humoral and T cell immunity to SARS-CoV-2 has been observed in healthy individuals^28^, and although the relative contributions of memory B cell and T cell responses have yet to be fully defined in SARS-CoV-2 immunity, the preservation of T cell immunity reported here should provide some reassurance for anti-TNF treated patients. However, it is noteworthy that one fifth made no anti-spike T cell response following two doses of either vaccine. Chronic TNF exposure, a feature of many IMIDs, can render T cells anergic and can be reversed by anti-TNF treatment^29^. This may in part explain why the magnitude of T cell responses observed in anti-TNF-treated patients in this study did not differ significantly from patients treated with vedolizumab.

## Limitations

Although our data show major differences in the magnitude and durability of antibody responses, we have not assessed the impact of biologic therapy on specific immunoglobulin classes, antibody neutralisation or mucosal immune responses, which may be impaired, in particular, with anti-a4b7 therapy^30,31^. However, previous studies have demonstrated that anti-RBD antibody levels such as the ones measured in this study, strongly correlate with Wuhan Hu-1 live virus neutralization assays^32^ and we have demonstrated here that early antibody responses to vaccination correlates with the subsequent risk of breakthrough infection in immunosuppressed patients.

## Conclusions

Infliximab was associated with attenuated, less durable vaccine induced anti-SARS-CoV-2 spike antibody responses and a 50% increase in subsequent breakthrough SARS-CoV-2 infection. Patients treated with anti-TNF therapy should be prioritised for third vaccine doses.

## Methods

### Patient and settings

impaCt of bioLogic therApy on saRs-cov-2 Infection and immuniTY (CLARITY) IBD is a UK wide, multicentre, prospective observational cohort study investigating the impact of infliximab and vedolizumab and/or concomitant immunomodulators (azathioprine, mercaptopurine, and methotrexate) on SARS-CoV-2 acquisition, illness, and immunity in patients with IBD.

Study methods have been previously described^10,11^. Consecutive patients were recruited at the time of attendance at infusion units between 22^nd^ September 2020 and 23^rd^ December 2020 (Supplementary Table 1). Patients aged 5 years and over, with a diagnosis of IBD, treated with infliximab or vedolizumab were eligible for inclusion. Follow-up visits coincided with biologic infusions and occurred eight-weekly. Here, we report vaccine-induced antibody responses after a second-dose of either the BNT162b2 or ChAdOx1 nCoV-19 vaccines. Participants were eligible for our primary immunogenicity analysis, if they had had an anti-S RBD antibody test between 14 and 70 days after a second-dose vaccine, defined as a second dose of any of the licenced COVID-19 vaccines, 10-14 weeks after the first dose. Anti-S RBD antibody levels were compared with samples from 605 fully vaccinated adult participants from the Virus Watch study, a household community cohort of 10,000 individuals representative of the UK population of England and Wales recruited between 1 June 2020 to 31 August 2021^19^. Peripheral blood mononuclear cells (PBMC) for T cell experiments were collected from patients 4 to 6 weeks after the first and second dose of vaccine at the time of biologic infusions, at selected sites which could facilitate PBMC extraction within 12 hours of venepuncture.

### Outcome measures

Our primary outcome was anti-S RBD antibodies 2 to 10 weeks after second dose of the BNT162b2 or ChAdOx1 nCoV-19 vaccines.

Secondary outcomes were:

i. the proportion of participants who seroconverted
ii. anti-spike T cell responses in patients following the first and second dose of vaccines (iii) the durability of vaccine responses
iii. risk of breakthrough infections two or more weeks after two doses of vaccine
iv. antibody concentrations and seroconversion rates in patients with PCR or serological evidence of past SARS-CoV-2 infection at, or prior, to the post-vaccination serum sample

### Variables

Variables recorded by participants were demographics (age, sex, ethnicity, comorbidities, height and weight, smoking status, and postcode), IBD disease activity (PRO2), SARS-CoV-2 symptoms aligned to the COVID-19 symptoms study (symptoms, previous testing, and hospital admissions for COVID-19), and vaccine uptake (type and date of primary vaccination). Study sites completed data relating to IBD history (age at diagnosis, disease duration, and phenotype according to the Montreal classifications, previous surgeries, and duration of current biologic and immunomodulator therapy)^10^. We linked our data by NHS number or Community Health Index to Public Health England, Scotland, and Wales who archive dates and results of all SARS-CoV-2 PCR tests undertaken and vaccines administered. Data were entered electronically into a purpose-designed REDCap database hosted at the Royal Devon and Exeter NHS Foundation Trust^33^. Participants without access to the internet or electronic device completed their questionnaires on paper case record forms that were subsequently entered by local research teams.

### Laboratory methods

To determine antibody responses specific to vaccination we used the Roche Elecsys Anti-SARS-CoV-2 spike (S) immunoassay^34^ alongside the nucleocapsid (N) immunoassay^35^. This double sandwich electrochemiluminescence immunoassay uses a recombinant protein of the receptor binding domain on the spike protein as an antigen for the determination of antibodies against SARS-CoV-2. Sample electrochemiluminescence signals are compared to an internal calibration curve and quantitative values are reported as units (U)/mL. In-house assay validation experiments were previously reported and included in the Supplementary methods^10,11^. Seroconversion was defined at a threshold of 15 U/mL. ElecSys Anti-SARS-CoV-2 spike (S) RBD concentrations of greater than or equal to 15 U/ml are associated with neutralization of ≥20% with a positive predictive value of 99.10 % (95% CI: 97.74-99.64)^11^.

At entry to CLARITY IBD and at follow-up visits, all patients were tested for previous SARS-CoV-2 infection using the Roche Elecsys anti-SARS-CoV-2 (N) immunoassay. We have previously reported that anti-N antibody responses following SARS-CoV-2 natural infection are impaired in patients treated with infliximab or vedolizumab^11^. As such, a threshold 0.12 times above the cut-off index was set, using receiver operator characteristic curve and area under the curve analysis of anti-N antibody results from participants two weeks following a PCR-confirmed infection to maximise specificity, beyond which patients were deemed to have had prior SARS-CoV-2 infection (Supplementary Figure 7). Patients with a PCR test confirming SARS-CoV-2 infection at any time prior to vaccination were deemed to have evidence of past infection irrespective of any antibody test result. Breakthrough infections were defined by a positive SARS-CoV-2 PCR test 2 or more weeks after the second vaccine dose.

### Peripheral blood mononuclear cell isolation

Whole blood was collected in lithium heparin tubes and PBMCs were isolated by density-gradient centrifugation using Lymphoprep™ (Stem Cell Technologies) layered on to SepMate™ (Stem Cell Technologies) tubes. PBMC isolation was performed within 12 hours of venepuncture. Purified PBMCs were cryopreserved in 10% DMSO/50% FBS and stored in liquid nitrogen pending batch analysis.

### Spike-peptide specific T cell responses

IFNg T cell ELISpot assays were performed using pre-coated plates (Mabtech 3420-2APT) and using the protocol described previously^28,32^. Two-hundred thousand cells were seeded per well and cells were stimulated with a peptide pool, containing 18 peptides derived from SARS-CoV-2 spike protein36 at a concentration of 10 μg/ml/peptide; the peptide pool utilises a mapped epitope pool (MEP) or 12-20mer peptides, mapped as eliciting high-prevalence CD4 responses covering diverse HLA-II haplotypes^28,32^. Use of this spike MEP in otherwise healthy SARS-CoV-2 seropositive individuals elicits a T cell response in 83% of individuals at 16 – 18 weeks after natural SARS-CoV-2 infection and 91% of healthy individuals 2-3 weeks after two dose vaccination with seronegative individuals showing a level of response indistinguishable from pre-pandemic controls^28,32^. Plates were cultured for 18-20 hours before development and data collected using an AID classic ELISpot plate reader (Autoimmun Diagnostika GMBH). Results are expressed as difference in (delta) spot forming cells (SFC) per 10^6^ PBMC between peptide stimulation and a media only control. A response below 2 standard deviations of the media only control wells was deemed to be a null response. Data was excluded if response to the positive control anti-CD3 stimulation was <200 SFC per 10^6^ PBMCs.

### Sample size

The sample size for CLARITY IBD was based on the number of participants required to demonstrate a difference in the impact of infliximab and vedolizumab on seroprevalence and seroconversion following SARS-CoV-2 infection, with an estimated background seroprevalence of 0.05. We calculated that a sample of 6970 patients would provide 80% power to detect differences in the seroprevalence of SARS-CoV-2 antibodies in infliximab-compared with vedolizumab-treated patients, whilst controlling for immunomodulator status at the 0.05 significance level.

## Statistical analyses

Analyses were undertaken using R 4.1.0 (R Foundation for Statistical Computing, Vienna, Austria). All tests were two tailed and p-values reported without correction for multiple testing. P-values <0.05 were considered significant. We included patients with missing clinical data in analyses for which they had data and have specified the denominator for each variable. Anti-S RBD antibody concentrations are reported as geometric means and standard deviations. Other continuous data are reported as median and interquartile range, and discrete data as numbers and percentages, unless otherwise stated.

Univariable analyses, using Spearman’s rank correlation coefficients, and t-tests of log-transformed anti-S RBD antibody concentration were used to identify demographic, disease, vaccine, and treatment-related factors associated with the concentration of anti-S RBD antibodies across the cohort. Crude sensitivity analyses excluding patients treated without a concomitant immunomodulator were undertaken to control for the effect of immunomodulator use on anti-S RBD antibody concentrations. Propensity matching was used to account for the other significant differences in baseline variables between infliximab- and vedolizumab-treated patients using the MatchIt package^37^. A priori, patients were matched exactly on diagnosis, immunomodulator use, and then using optimal matching, on age, number of comorbidities, ethnicity, and presence of active disease. Multivariable linear regression models were used to identify factors independently associated with log anti-S RBD concentration. A priori, we included age, ethnicity, biological medication and immunomodulator use. Results are presented after exponentiation, so that the coefficients of the model correspond to the fold change (FC) associated with each binary covariate. For age, a cut-off was chosen based on graphical inspection of the relationship between age and anti-S RBD antibody concentrations.

Mann-Whitney U test was used to compare the magnitude of T cell response (SFC/10^6^ PBMCs) stratified by treatment and vaccine received, and Spearman’s rank correlation coefficient was calculated to determine correlation between antibody and T cell responses.

Anti-S RBD antibody half-lives were estimated using an exponential model of decay. Linear mixed models were fit using the lme4 and lmerTest package, with biologic treatment and vaccine type as fixed effects and each subject as a random effect. Each of these effects were estimated independently for gradient and intercept. 95% confidence intervals of fixed effects were calculated using likelihood ratios. P values for comparison of half-lives were estimated from the full linear mixed effects model that incorporated vaccine, biologic drug and prior SARS-CoV-2 infection status.

We visualized durability of antibody responses by calculating 15-day rolling geometric mean anti-S RBD antibody concentrations. For this analysis we included participants who had an antibody test carried out between 1 and 70 days after second vaccine dose. Cox proportional hazard regression models were used to identify demographic, disease and treatment-related factors associated with the time to fall in anti-S RBD antibody concentration below the seroconversion threshold.

Kaplan-Meier curves and cox proportional hazard regression model was used to identify treatment-related factors associated with time to breakthrough infection. Linear regression model of log-transformed geometric mean anti-S RBD antibody concentration was used to determine the risk of breakthrough infections.

Where appropriate the same analyses were used to compare antibody responses in participants with PCR evidence of SARS-CoV-2 infection at any time prior to vaccination.

### Ethical consideration and role of funders

CLARITY IBD is an investigator-led, UK National Institute for Health Research COVID-19 urgent public health study, funded by the Royal Devon and Exeter NHS Foundation Trust, NIHR Imperial Biomedical Research Centre, Hull University Teaching Hospital NHS Trust, and by unrestricted educational grants from F. Hoffmann-La Roche AG (Switzerland), Biogen GmbH (Switzerland), Celltrion Healthcare (South Korea), Takeda (UK), and Galapagos NV (Belgium). None of our funding bodies had any role in study design, data collection or analysis, writing, or decision to submit for publication. Patients were included after providing informed, written consent. The sponsor was the Royal Devon and Exeter NHS Foundation Trust. The Surrey Borders Research Ethics committee approved the study (REC reference: REC 20/HRA/3114) in September 2020. The protocol is available online at https://www.clarityibd.org. The study was registered with the ISRCTN registry (ISRCTN45176516).

## Data Availability

The study protocol including the statistical analysis plan is available at www.clarityibd.org. Individual participant de-identified data that underlie the results reported in this article will be available immediately after publication for a period of 5 years. The data will be made available to investigators whose proposed use of the data has been approved by an independent review committee. Analyses will be restricted to the aims in the approved proposal. Proposals should be directed to. To gain access data requestors will need to sign a data access agreement.

## Code availability

Code used for data analysis will be available upon request directed to.

## Acknowledgements

CLARITY IBD is a UK National Institute for Health Research (NIHR) Urgent Public Health Study. The NIHR Clinical Research Network supported study set-up, site identification, and delivery of this study. This was facilitated by Professor Mark Hull, the National Speciality Lead for Gastroenterology. We acknowledge the contribution of our Patient Advisory Group who helped shape the trial design around patient priorities. Our partners, Crohn’s and Colitis UK (CCUK), continue to support this group and participate in Study Management Team meetings. We thank Professor Graham Cooke and Dr Katrina Pollock for their helpful discussions and review of the data. Laboratory tests were undertaken by the Exeter Blood Sciences Laboratory at the Royal Devon and Exeter NHS Foundation Trust. The Exeter NIHR Clinical Research Facility coordinated sample storage and management. Tariq Malik and James Thomas from Public Health England, Guy Stevens, Katie Donelon, Elen de Lacy from Public Health Wales and Johanna Bruce from Public Health Scotland supported linkage of central SARS-CoV-2 PCR test results with study data. Roche Diagnostics Limited provided the Elecsys Anti-SARS-CoV-2 immunoassay for the study. Faculty of Medicine at Imperial College London, Exeter NIHR Clinical Research Facility, Jeffrey Cheah Biomedical Centre at the University of Cambridge, Newcastle University Medical School and The Queen’s Medical Research Institute at the University of Edinburgh facilitated PBMC extractions for the T cell experiments. We thank Professor Robert Aldridge for access to data from the Virus Watch Collaborative. SL is supported by a Wellcome GW4-CAT fellowship. NC acknowledges support from CCUK. JCL is a Lister Prize Fellow and acknowledges support from the Cambridge NIHR Biomedical Research Centre and the Francis Crick Institute which receives core funding from Cancer Research UK (FC001169), the UK Medical Research Council (FC001169), and the Wellcome Trust (FC001169). GRJ is supported by a Wellcome Trust Clinical Research Career Development Fellowship (220725/Z/20/Z). CAL acknowledges support from the NIHR Newcastle Biomedical Research Centre and the support of the Programmed Investigation Unit at Royal Victoria Infirmary, Newcastle upon Tyne. CWL is funded by a UKRI Future Leaders Fellowship. RJB and DMA are supported by MRC (MR/W020610/1, MR/S019553/1, MR/R02622X/1 and⍰MR/V036939/1), NCSi4P, NIHR EME NIHR134607 and NIHR COV-LT2-0027,⍰Innovate UK SBRI894, NIHR Imperial Biomedical Research Centre (BRC):ITMAT, Cystic Fibrosis Trust SRC (2019SRC015), and Horizon 2020 Marie Skłodowska-Curie Innovative Training Network (ITN) European Training Network (No 860325). NP is supported by the NIHR Imperial Biomedical Research Center (BRC). We acknowledge the study co-ordinators of the Exeter Inflammatory Bowel Disease Research Group: Marian Parkinson and Helen Gardner-Thorpe for their ongoing administrative support to the study. The sponsor of the study was the Royal Devon and Exeter NHS Foundation Trust.

## Author Contributions

NAK, JRG, CB, SS, NP, TA participated in the conception and design of this study. CB was the project manager and coordinated patient recruitment. RN and TJM coordinated all biochemical analyses and central laboratory aspects of the project. SL, NAK, AS, DMS, CJR, RCS, SHK, FPP, KML, DKB, NC, DC, CB, MJ, SS, JLA, LC, JCL, CDM, ALH, PMI, GRJ, KBK, CAL, CWL, DMA, RJB, JRG, NP, TA were involved in the acquisition, analysis, or interpretation of data. DMS, CJR, KML, DKB, and FFP performed, analysed and interpreted T cell experiments. T cell experiments were supervised, designed, analysed and interpreted by RJB and DMA. Data analysis was done by NAK, DMS and RJB. Drafting of the manuscript was done by SL, NAK, NC, SS, CWL, DMA, RJB, JRG, NP, TA. NP and TA obtained the funding for the study. All the authors contributed to the critical review and final approval of the manuscript. NAK, NP and TA have verified the underlying data.

