## Supplementary information for "Antibody decay, T cell immunity and breakthrough infections following two SARS-CoV-2 vaccine doses in infliximab- and vedolizumab-treated patients"

### Supplementary Table 1: Contributors to the CLARITY IBD study

| Affiliation | First name | Surname |
|---|---|---|
| Barts Health NHS Trust | Klaartje | Kok |
| | Farjhana | Bokth |
| | Bessie | Cipriano |
| | Caroline | Francia |
| | Nosheen | Khalid |
| | Hafiza | Khatun |
| | Ashley | Kingston |
| | Irish | Lee |
| | Anouk | Lehmann |
| | Kinnari | Naik |
| | Elise | Pabriaga |
| | Nicolene | Plaatjies |
| | Kevin | Samuels |
| Barts Health NHS Trust (paediatric) | Bessie | Cipriano |
| | Kevin | Samuels |
| | Nicolene | Plaatjies |
| | Hafiza | Khatun |
| | Farjana | Bokth |
| | Elise | Pabriaga |
| | Caroline | Francia |
| Basingstoke and North Hampshire Hospital | Rebecca | Saich |
| | Hayley | Cousins |
| | Wendy | Fraser |
| | Rachel | Thomas |

| Affiliation | First name | Surname |
| --- | --- | --- |
| | Matthew | Brown |
| | Benjamin | White |
| Basingstoke and North Hampshire Hospital | Nikolaos. | Kirkineziadis |
| | Bernadette | Tilley |
| Birmingham Women's and Children's NHS Foundation Trust | Rafeeq | Muhammed |
| | Rehana | Bi |
| | Catherine | Cotter |
| | Jayne | Grove |
| | Kate | Hong |
| | Ruth | Howman |
| | Monica | Mitchell |
| | Sophie | Clayton |
| | Sugrah | Sultan |
| | Melanie | Rooney |
| | Charlotte | Cottrill |
| Bolton NHS Foundation Trust | Salil | Singh |
| | Chris | Dawe |
| | Robert | Hull |
| | Natalie | Silva |
| Borders General Hospital | Jonathan | Manning |
| | Lauren | Finlayson |
| | Allison | Roebuck |
| | Joy | Dawson |
| Calderdale and Huddersfield NHS Foundation Trust | Sunil | Sonwalkar |
| | Naomi | Chambers |
| | Matthew | Robinson |

| Affiliation | First name | Surname |
|---|---|---|
| | Andrew | Haigh |
| | Lear | Matapure |
| Cambridge University Hospitals NHS Foundation Trust | Tim | Raine |
| | Varun | George |
| | Christina | Kapizioni |
| | Konstantina | Strongili |
| | Tina | Thompson |
| | Mohamed | Ahmed |
| | Christos | Kontos |
| Cambridge Institute of Therapeutic Immunology and Infectious Disease, Jeffrey Cheah Biomedical Centre, University of Cambridge | Christophe | Bourges |
| | Isabella | Barbutti |
| | Megan E | Gozzard |
| Chelsea and Westminster Hospital NHS Foundation Trust | Philip | Hendy |
| | Rhian | Bull |
| | Patricia | Costa |
| | Lisa | Davey |
| | Hayley | Hannington |
| | Kribashnie | Nundlall |
| | Catarina | Martins |
| | Laura | Avanzi |
| | Jaime | Carungcong |
| | Sabrina | Barr |
| | Richard | Appleby |
| | Emma | Johnson |
| Chesterfield Royal Hospital | Kath | Phillis |
| | Rachel | Gascoyne |
| | Amanda | Crowder |

| Affiliation | First name | Surname |
|---|---|---|
| | Amanda | Whileman |
| Countess Of Chester Hospital NHS Foundation Trust | Ian | London |
| | Jenny | Grounds |
| | Emmeline | Martin |
| | Susie | Pajak |
| | Jude | Price |
| | Kathryn | Cawley |
| Darlington Memorial Hospital | Anjan | Dhar |
| | Ellen | Brown |
| | Amanda | Cowton |
| | Kimberley | Stamp |
| Dartford and Gravesham NHS Trust | Ben | Warner |
| | Carmel | Stuart |
| | Louise | Lacey |
| The Dudley Group NHS Foundation Trust | Shanika | de Silva |
| | Clare | Allcock |
| | Philip | Harvey |
| | Lesley | Jones |
| | Elise | Cooke |
| East and North Hertfordshire NHS Trust | Johanne | Brooks |
| | Pearl | Baker |
| | Hannah | Beadle |
| | Carina | Cruz |
| | Debbie | Potter |
| East Lancashire Hospitals NHS Trust | Joe | Collum |
| | Farzana | Masters |
| Glangwili Hospital | Aashish | Kumar |

| Affiliation | First name | Surname |
|---|---|---|
| | Samantha | Coetzee |
| | Mihaela | Peiu |
| | Becky | Icke |
| | Meena | Raj |
| Great Ormond Street Hospital | Edward | Gaynor |
| | Sibongile | Chadokufa |
| | Bonita | Huggett |
| | Hamza | Meghari |
| | Sara | El-Khouly |
| | Fevronia | Kiparissi |
| | Waffa | Girshab |
| Great Western Hospitals NHS Foundation Trust | Andrew | Claridge |
| | Emily | Fowler |
| | Laura | McCafferty |
| Guy's and St Thomas' NHS Foundation Trust | Peter | Irving |
| | Karolina | Christodoulides |
| | Angela | Clifford |
| | Patrick | Dawson |
| | Sailish | Honap |
| | Samuel | Lim |
| | Raphael | Luber |
| | Karina | Mahiouz |
| | Susanna | Meade |
| | Parizade | Raymode |
| | Rebecca | Reynolds |
| | Anna | Stanton |
| | Sherill | Tripoli |

| Affiliation | First name | Surname |
|---|---|---|
| | Naomi | Hare |
| | Senthuran | Balachandran |
| | Emma | North |
| | Jessica | North |
| | Bria | Browne |
| | Ella | Jameson |
| The Hillingdon Hospitals NHS Foundation Trust | Yih Harn | Siaw |
| | Lane | Manzano |
| | Jonathan | Segal |
| | Ibrahim | Al-Bakir |
| | Imran | Khakoo |
| Homerton University Hospital Foundation Trust | Nora | Thoua |
| | Katherine | Davidson |
| | Jagrul | Miah |
| | Lisa | Canclini |
| | Alex | Hall |
| Hull University Teaching Hospitals NHS Trust | Shaji | Sebastian |
| | Melony | Hayes |
| | Sally | Myers |
| | Alison | Talbot |
| | Jack | Turnbull |
| | Emma | Whitehead |
| | Katie | Stamp |
| | Alison | Pattinson |
| | Verghese | Mathew |
| | Leanne | Sherris |

| Affiliation | First name | Surname |
|---|---|---|
| Hull University Teaching Hospitals NHS Trust | Angela | Harvey |
| Imperial College Healthcare NHS Trust | Lucy | Hicks |
| | Tara-Marie | Byrne |
| | Leilani | Cabreros |
| | Hannah | Downing-Wood |
| | Sophie | Hunter |
| | Mohammad Aamir | Saifuddin |
| | Hemanth | Prabhudev |
| | Sharmili | Balarajah |
| Faculty of Medicine, Imperial College London | Hajir | Ibraheim |
| | Melissa | Torkizadeh |
| | Jonathan W | Lo |
| | Zhigang | Liu |
| James Paget University Hospitals NHS Foundation Trust | Helen | Sutherland |
| | Elva | Wilhelmsen |
| | Katherine | Mackintosh |
| Kettering General Hospital | Ajay M | Verma |
| | Juliemol | Sebastian |
| | Mohammad Farhad | Peerally |
| | Parizade | Raymode |
| | Anne-marie | Guerdette |
| King's College Hospital NHS Foundation Trust | Alexandra | Kent |
| | Lee Meng | Choong |
| | Benedetta | Pantaloni |
| | Pantelis | Ravdas |
| King's College Hospital NHS Foundation Trust (paediatric) | Babu | Vadamalayan |

| Affiliation | First name | Surname |
|---|---|---|
| King’s Mill Hospital | Stephen | Foley |
| | Becky | Arnold |
| | Cheryl | Heeley |
| | Wayne | Lovegrove |
| | Donna | Sowton |
| | Lynne | Allsop |
| | Heidi | Gregory |
| Liverpool University Hospitals NHS Foundation Trust | Philip J | Smith |
| | Giovanna | Bretland |
| | Sarah | King |
| | Martina | Lofthouse |
| | Lindsey | Rigby |
| | Sreedhar | Subramanian |
| | David | Tyrer |
| | Kate | Martin |
| | Christopher | Probert |
| London North West University Healthcare NHS Trust | Nikolaos | Kamperidis |
| | Temi | Adedoyin |
| | Manisha | Baden |
| | Jeannette | Brown |
| | Feba | Chacko |
| | Michela | Cicchetti |
| | Mohammad | Saifuddin |
| | Priya | Yesupatham |
| Maidstone and Tunbridge Wells NHS Trust | Rohit | Gowda |
| | Maureen | Williams |
| Manchester University NHS Foundation | Karen | Kemp |

| Affiliation | First name | Surname |
|---|---|---|
| Trust | Rima | Akhand |
| | Glaxy | Gray |
| | Anu | John |
| | Maya | John |
| | Tasnim | Mohammed |
| | Diamond | Sathe |
| | Natasha | Jones |
| | Jennifer | Soren |
| The Mid Yorkshire Hospitals NHS Trust | Michael | Sprakes |
| | Julie | Burton |
| | Patricia | Kane |
| | Stephanie | Lupton |
| | Jacqueline | Bartholomew |
| Milton Keynes University Hospital | George | MacFaul |
| | Diane | Scaletta |
| | Loria | Siamia |
| | Felicity | Williams |
| | Chloe | Green |
| | Zeljka | Ver |
| Newcastle Hospitals NHS Foundation Trust | Chris | Lamb |
| | Mary | Doona |
| | Ashleigh | Hogg |
| | Lesley | Jeffrey |
| | Andrew | King |
| | R Alexander | Speight |
| | Jennifer | Doyle |
| | Ruth | Owen |

| Affiliation | First name | Surname |
|---|---|---|
| Ninewells Hospital & Medical School | Craig | Mowat |
| | Debbie | Rice |
| | Susan | MacFarlane |
| | Anne | MacLeod |
| | Samera | Mohammed |
| Ninewells Hospital & Medical | Shona | Murray |
| | Anne | Elliott |
| Norfolk and Norwich University Hospitals NHS Foundation Trust | Mary | Anne Morris |
| | Louise | Coke |
| | Grace | Hindle |
| | Eirini | Kolokouri |
| | Catherine | Wright |
| | Claire | Lee |
| | Nicola | Ward |
| | Adele | Dann |
| North Bristol NHS Trust | Melanie | Lockett |
| | Charlotte | Cranfield |
| | Louise | Jennings |
| | Ankur | Srivastava |
| | Lana | Ward |
| | Nouf | Jeynes |
| | Poonam | Ranga |
| North Tyneside General Hospital | Praveen | Rajasekhar |
| | Lisa | Gallagher |
| | Linda | Patterson |
| | Jill | Ward |
| | Rae | Basnett |

| Affiliation | First name | Surname |
|---|---|---|
| | Judy | Murphy |
| | Lauren | Parking |
| | Emma | Lawson |
| | Stacey | Short |
| Nottingham University Hospitals NHS Trust | David | Devadason |
| | Gordon | Moran |
| | Neelam | Khan |
| | Lauren | Tarr |
| | Charmaine | Olivia |
| The Pennine Acute Hospitals NHS Trust | Jimmy | Limdi |
| | Kay | Goulden |
| | Asad | Javed |
| | Lauren | McKenzie |
| Portsmouth Hospitals NHS Trust | Pradeep | Bhandari |
| | Michelle | Baker-Moffatt |
| | Joanne | Dash |
| | Allison | Le Poidevin |
| | Hayley | Downe |
| | Lucille | Bombeo |
| | Helen | Blackman |
| The Queen Elizabeth Hospital Kings Lynn NHS Trust | Alan | Wiles |
| | Hannah | Bloxham |
| | Jose | Dias |
| | Evelyn | Nadar |
| | Hollie. | Curgenven |
| Queen Elizabeth University Hospital, Glasgow | Jonathan | Macdonald |
| | Shona | Finan |

| Affiliation | First name | Surname |
|---|---|---|
| | Faye | McMeeken |
| | Misbah | Mahmood |
| | Stephanie | Shields |
| | John Paul | Seenan |
| Royal Berkshire NHS Foundation Trust | Des | DeSilva |
| | Susanna | Malkakorpi |
| | Rachel | Carson |
| University Hospitals Dorset NHS Foundation Trust | Simon | Whiteoak |
| | Kelli | Edger-Earley |
| | Luke | Vamplew |
| Royal Cornwall Hospitals NHS Trust | Sarah | Ingram |
| | Sharon | Botfield |
| | Fiona | Hammonds |
| | Clare | James |
| Royal Devon and Exeter NHS Foundation Trust | Tariq | Ahmad |
| | Gemma | Aspinall |
| | Sarah | Hawkins |
| | Suzie | Marriott |
| | Clare | Redstone |
| | Halina | Windak |
| | Ana-Marie | Adam |
| | Hannah | Mabb |
| Royal Free London NHS Foundation Trust | Charles | Murray |
| | Cynthia | Diaba |
| | Fexy | Joseph |
| | Glykeria | Pakou |
| | Yvonne | Gleeson |

| Affiliation | First name | Surname |
|---|---|---|
| Royal Glamorgan Hospital | James | Berrill |
| | Natalie | Stroud |
| | Carla | Pothecary |
| | Lisa | Roche |
| | Keri | Turner |
| | Lisa | Deering |
| | Lynda | Israel |
| Royal Gwent Hospital | Evelyn | Baker |
| | Sean | Cutler |
| | Rina | Mardania Evans |
| | Maxine | Nash |
| | Georgina | Mallison |
| | Anna | Roynon |
| Royal Hampshire County Hospital | John | Gordon |
| | Emma | Levell |
| | Silvia | Zagalo |
| | Wendy | Fraser |
| | Ina | Hoad |
| | Nikolaos | Kirkineziadis |
| Royal Hospital for Sick Children, Edinburgh | Richard | Russell |
| | Paul | Henderson |
| | Margaret | Millar |
| Royal Manchester Children's Hospital | Andrew | Fagbemi |
| | Felicia | Jennings |
| | Imelda | Mayor |
| | Jill | Wilson |
| Royal Surrey County Hospital | Christopher | Alexakis |

| Affiliation | First name | Surname |
|---|---|---|
| | Natalia | Michalak |
| Royal United Hospitals Bath | John | Saunders |
| | Helen | Burton |
| | Vanessa | Cambridge |
| | Tonia | Clark |
| | Charlotte | Ekblad |
| | Sarah | Hierons |
| | Joyce | Katebe |
| | Emma | Saunsbury |
| | Rachel | Perry |
| The Royal Wolverhampton NHS Trust | Matthew | Brookes |
| | Kathryn | Davies |
| | Marie | Green |
| | Ann | Plumbe |
| Salford Royal NHS Foundation Trust | Clare | Ormerod |
| | Helen | Christensen |
| | Anne | Keen |
| | Jonathan | Ogor |
| Salisbury District Hospital | Alpha | Anthony |
| | Emily | Newitt |
| | Fiona | Trim |
| | Ruth | Casey |
| | Katherine | Seymour |
| Sandwell and West Birmingham NHS Trust | Edward | Fogden |
| | Kalisha | Russell |
| Sheffield Teaching Hospitals NHS Foundation Trust | Anne | Phillips |
| | Muaad | Abdulla |

| Affiliation | First name | Surname |
|---|---|---|
| Shrewsbury and Telford Hospital NHS Trust | Jeff | Butterworth |
| | Colene | Adams |
| | Elizabeth | Buckingham |
| | Danielle | Childs |
| | Alison | Magness |
| | Jo | Stickley |
| | Nichola | Motherwell |
| | Louise | Tonks |
| | Hannah | Gibson |
| | S | Pajak |
| Singleton Hospital | Caradog | Thomas |
| | Elaine | Brinkworth |
| | Lynda | Connor |
| | Amanda | Cook |
| | Tabitha | Rees |
| | Rachel | Harford |
| Somerset NHS Foundation Trust | Emma | Wesley |
| | Alison | Moss |
| | Jacob | Lucas |
| | Claire | Lorimer |
| | Maria | Oleary |
| | Maxine | Dixon |
| South Tees Hospitals NHS Foundation Trust | Arvind | Ramadas |
| | Julie | Tregonning |
| | Olaku | Okeke |
| | Wendy | Jackson |
| Southend University Hospital NHS | Ioannis | Koumoutsos |

| Affiliation | First name | Surname |
|---|---|---|
| Foundation Trust | Viji | George |
| | Swapna | Kunhunny |
| | Sophie | Laverick |
| | Isla | Anderson |
| | Sophie | Smith |
| St George's University Hospitals NHS Foundation Trust | Kamal | Patel |
| | Mariam | Ali |
| | Hilda | Mhandu |
| | Aleem | Rana |
| | Katherine | Spears |
| | Joana | Teixeira |
| | Richard | Pollok |
| | Mark | Mencias |
| | Abigail | Seaward |
| | Jessica | Sousa |
| St Georges University Hospital NHS Trust | Nooria | Said |
| | Mark | Soomaroo |
| | Valentina | Raspa |
| | Asha | Tacouri |
| St George's University Hospitals NHS Foundation Trust (paediatric) | Nicholas | Reps |
| | Rebecca | Martin |
| St James's University Hospital | Christian | Selinger |
| | Jenelyn | Carbonell |
| | Felicia | Onovira |
| | Doris | Quartey |
| | Alice | L'Anson |
| | Andrew | Ashworth |

| Affiliation | First name | Surname |
|---|---|---|
| | Jessica | Bailey |
| | Angie | Dunn |
| Stockport NHS Foundation Trust | Zahid | Mahmood |
| | Racheal | Campbell |
| | Liane | Marsh |
| Surrey and Sussex Healthcare NHS Trust | Monira | Rahman |
| | Sarah | Davies |
| | Ruth | Habibi |
| | Ellen | Jessup-Dunton |
| | Teishel | Joefield |
| | Reina | Layug |
| Tameside and Glossop Integrated Care NHS Foundation Trust | Vinod | Patel |
| | Joanne | Vere |
| | Victoria | Turner |
| | Susan | Kilroy |
| Torbay and South Devon NHS Foundation Trust | Gareth | Walker |
| | Stacey | Atkins |
| | Jasmine | Growdon |
| | Charlotte | McNeill |
| University Hospitals Birmingham NHS Foundation Trust | Rachel | Cooney |
| | Lillie | Bennett |
| | Louise | Bowlas |
| | Sharafaath | Shariff |
| University Hospitals Bristol NHS Foundation Trust | Aileen | Fraser |
| | Dwayne | Punnette |
| | Charlotte | Bishop-Hurman |
| | Elizabeth | Undrell |

| Affiliation | First name | Surname |
|---|---|---|
| | Katherine | Belfield |
| University Hospitals of Derby and Burton NHS Foundation Trust | Said | Din |
| | Catherine | Addleton |
| | Marie | Appleby |
| | Johanna | Brown |
| | Kathleen | Holding |
| University Hospitals of Leicester NHS Trust | Patricia | Hooper |
| | John | deCaestecker |
| | Olivia | Watchorn |
| University Hospitals Plymouth NHS Trust | Chris | Hayward |
| | Susan | Inniss |
| | Lucy | Pritchard |
| | Karen | Rudge |
| | Amanda | Carney |
| United Lincolnshire Hospitals NHS Trust | Jervoise | Andreyev |
| | Caroline | Hayhurst |
| | Carol | Lockwood |
| | Lynn | Osborne |
| | Amanda | Roper |
| | Karen | Warner |
| | Julia | Hindle |
| | Caroline | Watt |
| | Kinga | Szymiczek |
| University College London Hospitals NHS Foundation Trust | Shameer | Mehta |
| | James | Bell |
| | William | Blad |
| | Lisa | Whitley |

| Affiliation | First name | Surname |
|---|---|---|
| University Hospital Llandough | Durai | Dhamaraj |
| | Mark | Baker |
| | Elizabeth | John Sivamurugan |
| | Mim | Evans |
| University Hospital Southampton NHS Foundation Trust | Fraser | Cummings |
| | Clare | Harris |
| | Amy | Jones |
| | Liga | Krauze |
| | Sohail | Rahmany |
| | Michelle | Earl |
| | Jenny | Vowles |
| | Audrey | Torokwa |
| | Mirela | Petrova |
| | Andrew | Procter |
| | Jo | Stanley |
| | Claudia | Silvamoniz |
| | Marion | Bettey |
| University Hospital of Wales (paediatric) | Amar | Wahid |
| | Zoe | Morrison |
| | Rhian | Thomas-Turner |
| | Louise | Yendle |
| | Jennifer | Muller |
| Exeter NIHR Clinical Research Facility, University of Exeter | Marcus | Mitchell |
| | John | Kirkwood |
| | Anna | Barnes |
| West Hertfordshire Hospitals NHS Trust | Rakesh | Chaudhary |
| | Melanie | Claridge |

| Affiliation | First name | Surname |
|---|---|---|
| | Chiara | Ellis |
| | Cheryl | Kemp |
| | Ogwa | Tobi |
| | Jentus | Milton |
| West Middlesex University Hospital | Emma | Johnston |
| | Metod | Oblak |
| | Richard | Appleby |
| West Suffolk NHS Foundation Trust | Jo | Godden |
| Western General Hospital | Charlie | Lees |
| | Debbie | Alexander |
| | Kate | Covil |
| | Lauranne | Derikx |
| | Sryros | Siakavellas |
| | Helen | Baxter |
| | Scott | Robertson |
| | Linda | Smith |
| | Beena | Poulose |
| | Anne | Colemam |
| | Margareta | Balint |
| | Gareth | Rhys-Jones |
| Withybush General Hospital | Kerrie | Johns |
| | Rachel | Hughes |
| | Janet | Phipps |
| | Abigail | Taylor |
| Withybush General Hospital | Catherine | MacPhee |
| | Suzanne | Brooks |
| Yeovil District Hospital NHS Foundation | Katie | Smith |

| Affiliation | First name | Surname |
|---|---|---|
| Trust | Linda | Howard |
| | Dianne | Wood |
| York Teaching Hospital NHS Foundation Trust | Ajay | Muddu |
| | Laura | Barman |
| | Janine | Mallinson |
| | Tania | Neale |
| | Diana | Ionita |
| | Kerry | Elliot |
| | Alison | Turnball |
| Ysbyty Gwynedd | Iola | Thomas |
| | Kelly | Andrews |
| | Jonathon | Sutton |
| | Caroline | Mulvaney Jones |
| | Julia | Roberts |
| | Jeannie | Bishop |

#### Supplementary Table 2: Baseline characteristics of unique participants included in T cell studies stratified by biologic treatment at time of first vaccine dose

| Variable | Level | Vedolizumab | Infliximab | Overall | p |
|---|---|---|---|---|---|
| Vaccine | BNT162b2 | 49.3% (35/71) | 49.3% (104/211) | 49.3% (139/282) | 1.0 |
| | ChAdOx1 nCoV-19 | 50.7% (36/71) | 50.7% (107/211) | 50.7% (143/282) | |
| Age (years) | | 45.1 (34.0 - 54.5) | 38.1 (30.0 - 48.9) | 39.8 (30.9 - 49.7) | 0.0012 |
| Sex | Female | 47.9% (34/71) | 39.8% (84/211) | 41.8% (118/282) | 0.27 |
| | Male | 52.1% (37/71) | 60.2% (127/211) | 58.2% (164/282) | |
| | Intersex | 0.0% (0/71) | 0.0% (0/211) | 0.0% (0/282) | |
| | Prefer not to say | 0.0% (0/71) | 0.0% (0/211) | 0.0% (0/282) | |
| Ethnicity | White | 80.3% (57/71) | 82.5% (174/211) | 81.9% (231/282) | 0.83 |
| | Asian | 15.5% (11/71) | 11.8% (25/211) | 12.8% (36/282) | |
| | Mixed | 1.4% (1/71) | 1.4% (3/211) | 1.4% (4/282) | |
| | Black | 2.8% (2/71) | 2.4% (5/211) | 2.5% (7/282) | |
| | Other | 0.0% (0/71) | 1.9% (4/211) | 1.4% (4/282) | |
| Diagnosis | Crohn's disease | 28.2% (20/71) | 67.8% (143/211) | 57.8% (163/282) | <0.0001 |
| | UC/IBDU | 71.8% (51/71) | 32.2% (68/211) | 42.2% (119/282) | |
| Duration of IBD (years) | | 11.0 (6.5 - 21.0) | 10.0 (5.0 - 18.0) | 10.0 (5.0 - 19.0) | 0.12 |
| Age at IBD diagnosis (years) | | 27.8 (20.2 - 39.4) | 24.8 (17.2 - 34.0) | 25.8 (18.8 - 35.3) | 0.023 |
| Immunomodulators at vaccine | | 10.1% (7/69) | 62.6% (132/211) | 49.6% (139/280) | <0.0001 |
| 5-ASA | | 31.9% (22/69) | 19.4% (41/211) | 22.5% (63/280) | 0.045 |
| Steroids | | 7.2% (5/69) | 2.4% (5/211) | 3.6% (10/280) | 0.070 |
| BMI | | 25.6 (22.0 - 28.9) | 24.6 (21.8 - 27.1) | 24.7 (21.9 - 27.4) | 0.38 |
| Heart disease | | 2.8% (2/71) | 1.9% (4/211) | 2.1% (6/282) | 0.64 |
| Diabetes | | 9.9% (7/71) | 3.3% (7/211) | 5.0% (14/282) | 0.051 |
| Lung disease | | 21.1% (15/71) | 13.7% (29/211) | 15.6% (44/282) | 0.18 |
| Kidney disease | | 4.2% (3/71) | 0.9% (2/211) | 1.8% (5/282) | 0.10 |
| Cancer | | 2.8% (2/71) | 0.0% (0/211) | 0.7% (2/282) | 0.063 |
| Smoker | Yes | 4.2% (3/71) | 9.0% (19/211) | 7.8% (22/282) | 0.028 |
| | Not currently | 38.0% (27/71) | 22.3% (47/211) | 26.2% (74/282) | |
| | Never | 57.7% (41/71) | 68.7% (145/211) | 66.0% (186/282) | |
| Exposure to documented cases of COVID-19 | | 8.5% (6/71) | 4.3% (9/210) | 5.3% (15/281) | 0.22 |
| Income deprivation score | | 0.092 (0.048 - 0.163) | 0.084 (0.046 - 0.140) | 0.086 (0.046 - 0.146) | 0.26 |
| Active disease (PRO2) | | 6.2% (4/65) | 2.5% (5/201) | 3.4% (9/266) | 0.23 |
| Time between vaccine doses (weeks) | | 10.9 (10.0 - 11.1) | 10.7 (9.9 - 11.2) | 10.9 (9.9 - 11.1) | 0.35 |

#### Supplementary Table 3: Baseline characteristics of participants who had a SARS-CoV-2 infection prior to receiving 2 doses of a SARS-CoV-2 vaccine

| Variable | Level | Vedolizumab | Infliximab | p |
|---|---|---|---|---|
| Vaccine | BNT162b2 | 37.5% (84/224) | 41.3% (217/525) | 0.37 |
| | ChAdOx1 nCoV-19 | 62.5% (140/224) | 58.7% (308/525) | |
| Peak anti-N antibody concentration prior to first dose (U/mL) | | 0.9 (14.2) | 0.3 (5.4) | <0.0001 |
| Peak anti-S RBD antibody concentration prior to first dose (U/mL) | | 4.8 (20.9) | 1.3 (7.2) | <0.0001 |
| Peak anti-N antibody concentration prior to second dose (U/mL) | | 2.7 (16.5) | 0.6 (6.4) | <0.0001 |
| SARS-CoV-2 infection mode of diagnosis | Positive SARS-CoV-2 PCR test | 34.8% (78/224) | 31.0% (163/525) | 0.35 |
| | Anti-N antibody concentration ≥0.12 (U/mL) prior to second dose | 97.3% (218/224) | 95.8% (503/525) | 0.40 |
| | Positive SARS-CoV-2 PCR test and anti-N antibody concentration ≥0.12 (U/mL) prior to second dose | 34.8% (78/224) | 31.0% (163/525) | 0.35 |
| Age (years) | | 43.4 (34.3 - 56.3) | 38.8 (29.6 - 51.7) | <0.0001 |
| Sex | Female | 53.1% (119/224) | 43.8% (230/525) | 0.036 |
| | Male | 46.9% (105/224) | 56.0% (294/525) | |
| | Intersex | 0.0% (0/224) | 0.0% (0/525) | |
| | Prefer not to say | 0.0% (0/224) | 0.2% (1/525) | |
| Ethnicity | White | 88.8% (198/223) | 85.9% (451/525) | 0.86 |
| | Asian | 7.6% (17/223) | 8.4% (44/525) | |
| | Mixed | 1.3% (3/223) | 2.3% (12/525) | |
| | Black | 1.8% (4/223) | 2.9% (15/525) | |
| | Other | 0.4% (1/223) | 0.6% (3/525) | |
| Diagnosis | Crohn's disease | 32.6% (73/224) | 66.3% (348/525) | <0.0001 |
| | UC/IBDU | 67.4% (151/224) | 33.7% (177/525) | |
| Duration of IBD (years) | | 9.0 (5.0 - 16.0) | 8.0 (3.0 - 15.0) | 0.032 |
| Age at IBD diagnosis (years) | | 31.4 (23.1 - 42.8) | 27.6 (20.3 - 37.9) | 0.0014 |
| Immunomodulators at vaccine | | 17.2% (38/221) | 57.4% (300/523) | <0.0001 |
| 5-ASA | | 35.3% (78/221) | 22.2% (116/523) | 0.00025 |
| Steroids | | 5.0% (11/221) | 3.3% (17/523) | 0.29 |
| BMI | | 27.1 (24.2 - 31.8) | 25.8 (23.4 - 30.1) | 0.0082 |
| Heart disease | | 4.5% (10/222) | 2.5% (13/524) | 0.17 |
| Diabetes | | 5.8% (13/223) | 3.8% (20/525) | 0.24 |
| Lung disease | | 19.7% (44/223) | 10.7% (56/525) | 0.0014 |
| Kidney disease | | 1.8% (4/223) | 0.6% (3/525) | 0.21 |
| Cancer | | 1.3% (3/223) | 0.4% (2/525) | 0.16 |
| Smoker | Yes | 7.6% (17/223) | 10.9% (57/525) | 0.15 |
| | Not currently | 35.0% (78/223) | 29.0% (152/525) | |
| | Never | 57.4% (128/223) | 60.2% (316/525) | |
| Exposure to documented cases of COVID-19 | | 19.6% (44/224) | 18.1% (95/524) | 0.68 |
| Income deprivation score | | 0.102 (0.058 - 0.178) | 0.103 (0.057 - 0.185) | 0.99 |
| Active disease (PRO2) | | 6.2% (13/209) | 5.5% (27/492) | 0.72 |
| Time between vaccine doses (weeks) | | 11.0 (9.9 - 11.6) | 11.0 (10.0 - 11.3) | 0.66 |
| Time from second dose to serum sample (weeks) | | 5.6 (3.9 - 7.6) | 5.4 (3.6 - 7.7) | 0.81 |

Abbreviations: anti-N = anti-nucleocapsid; anti-S RBD = anti-spike receptor binding domain; IBD = inflammatory bowel disease; 5-ASA = 5-aminosalicylic acid; BMI = Body Mass Index; PRO2 = IBD disease activity. Values presented are median (interquartile range) or percentage (numerator/denominator). P values represent the results of a Mann Whitney U, Kruskal Wallis or Fisher's exact test.

#### Supplementary Table 4: Baseline characteristics of participants following propensity matching of baseline variables

| Variable | Level | Infliximab | Vedolizumab | p |
|---|---|---|---|---|
| Vaccine | BNT162b2 | 42.5% (329/774) | 40.4% (313/774) | 0.44 |
| | ChAdOx1 nCoV-19 | 57.5% (445/774) | 59.6% (461/774) | |
| Age (years) | | 46.5 (34.6 - 59.4) | 46.5 (34.6 - 59.5) | 0.91 |
| Sex | Female | 48.0% (373/777) | 49.2% (382/776) | 0.40 |
| | Male | 52.0% (404/777) | 50.5% (392/776) | |
| | Prefer not to say | 0.0% (0/777) | 0.3% (2/776) | |
| Ethnicity | White | 91.5% (711/777) | 89.9% (698/776) | 0.82 |
| | Asian | 5.3% (41/777) | 6.2% (48/776) | |
| | Mixed | 1.9% (15/777) | 2.4% (19/776) | |
| | Black | 0.6% (5/777) | 0.5% (4/776) | |
| | Other | 0.6% (5/777) | 0.9% (7/776) | |
| Diagnosis | Crohn's disease | 41.6% (323/777) | 41.6% (323/776) | 1.0 |
| | Ulcerative colitis | 55.2% (429/777) | 55.2% (428/776) | |
| | IBD-unclassified | 3.2% (25/777) | 3.2% (25/776) | |
| Duration of IBD (years) | | 9.0 (4.0 - 17.0) | 9.0 (4.0 - 16.0) | 0.79 |
| Age at IBD diagnosis (years) | | 31.8 (22.6 - 46.3) | 32.7 (22.7 - 45.4) | 0.63 |
| Immunomodulators at recruitment | | 25.6% (199/777) | 25.5% (198/776) | 1.0 |
| 5-ASA at recruitment | | 31.4% (242/770) | 32.3% (247/765) | 0.74 |
| Steroids in 2020 | | 3.9% (30/770) | 5.4% (41/765) | 0.18 |
| BMI | | 26.6 (23.4 - 29.9) | 25.9 (23.1 - 29.7) | 0.12 |
| Heart disease | | 4.4% (34/777) | 3.5% (27/776) | 0.43 |
| Diabetes | | 5.1% (40/777) | 7.0% (54/776) | 0.14 |
| Lung disease | | 15.1% (117/777) | 15.5% (120/776) | 0.83 |
| Kidney disease | | 1.2% (9/777) | 1.5% (12/776) | 0.52 |
| Cancer | | 0.4% (3/777) | 0.9% (7/776) | 0.22 |
| Smoker | Yes | 9.8% (76/777) | 8.6% (67/776) | 0.62 |
| | Not currently | 34.4% (267/777) | 36.3% (282/776) | |
| | Never | 55.9% (434/777) | 55.0% (427/776) | |
| Exposure to documented cases of COVID-19 | | 9.1% (71/777) | 7.3% (57/776) | 0.23 |
| Income deprivation score | | 0.094 (0.054 - 0.154) | 0.089 (0.053 - 0.147) | 0.65 |
| Active disease (PRO2) | | 10.2% (79/777) | 8.8% (68/776) | 0.39 |

Abbreviations: IBD = inflammatory bowel disease; 5-ASA = 5-aminosalicylic acid; BMI = Body Mass Index; PRO2 = IBD disease activity. Values presented are median (interquartile range) or percentage (numerator/denominator). P values represent the results of a Mann Whitney U, Kruskal Wallis or Fisher’s exact test.

Supplementary Table 5: Summary statistics for each interaction term in the linear mixed model used to estimate anti-S RBD antibody half-life. Modification of antibody half-life by biologic type, vaccine type and history of prior SARS-CoV-2 infection was evaluated by adding interaction terms in the model.

| **Type** | **Biologic: Infliximab** | **Vaccine: BNT162b2** | **Prior infection** | **Estimate (95% CI)** | **P value** |
|---|---|---|---|---|---|
| Intercept | | | | 9.81 (9.63, 9.99) | <0.0001 |
| Intercept | ✓ | | | -1.87 (-2.09, -1.66) | <0.0001 |
| Intercept | | ✓ | | 2.80 (2.51, 3.08) | <0.0001 |
| Intercept | | | ✓ | 2.04 (1.65, 2.43) | <0.0001 |
| Intercept | ✓ | ✓ | | -0.98 (-1.32, -0.64) | <0.0001 |
| Intercept | ✓ | | ✓ | 0.07 (-0.39, 0.54) | 0.75 |
| Intercept | | ✓ | ✓ | -1.41 (-2.04, -0.79) | <0.0001 |
| Intercept | ✓ | ✓ | ✓ | 1.31 (0.58, 2.05) | 0.00046 |
| Gradient | | | | -0.106 (-0.118, -0.094) | <0.0001 |
| Gradient | ✓ | | | -0.083 (-0.098, -0.068) | <0.0001 |
| Gradient | | ✓ | | -0.034 (-0.052, -0.016) | 0.00021 |
| Gradient | | | ✓ | 0.032 (0.005, 0.059) | 0.019 |
| Gradient | ✓ | ✓ | | -0.033 (-0.055, -0.011) | 0.0028 |
| Gradient | ✓ | | ✓ | 0.068 (0.036, 0.100) | <0.0001 |
| Gradient | | ✓ | ✓ | 0.005 (-0.035, 0.046) | 0.81 |
| Gradient | ✓ | ✓ | ✓ | 0.015 (-0.033, 0.062) | 0.55 |

Supplementary Figure 1: Proportion of patients with an anti-S RBD antibody concentration greater than the seroconversion threshold of 15U/mL at 2-10 weeks following two doses of SARS-CoV-2 vaccine stratified by biologic treatment, immunomodulator status and vaccine type

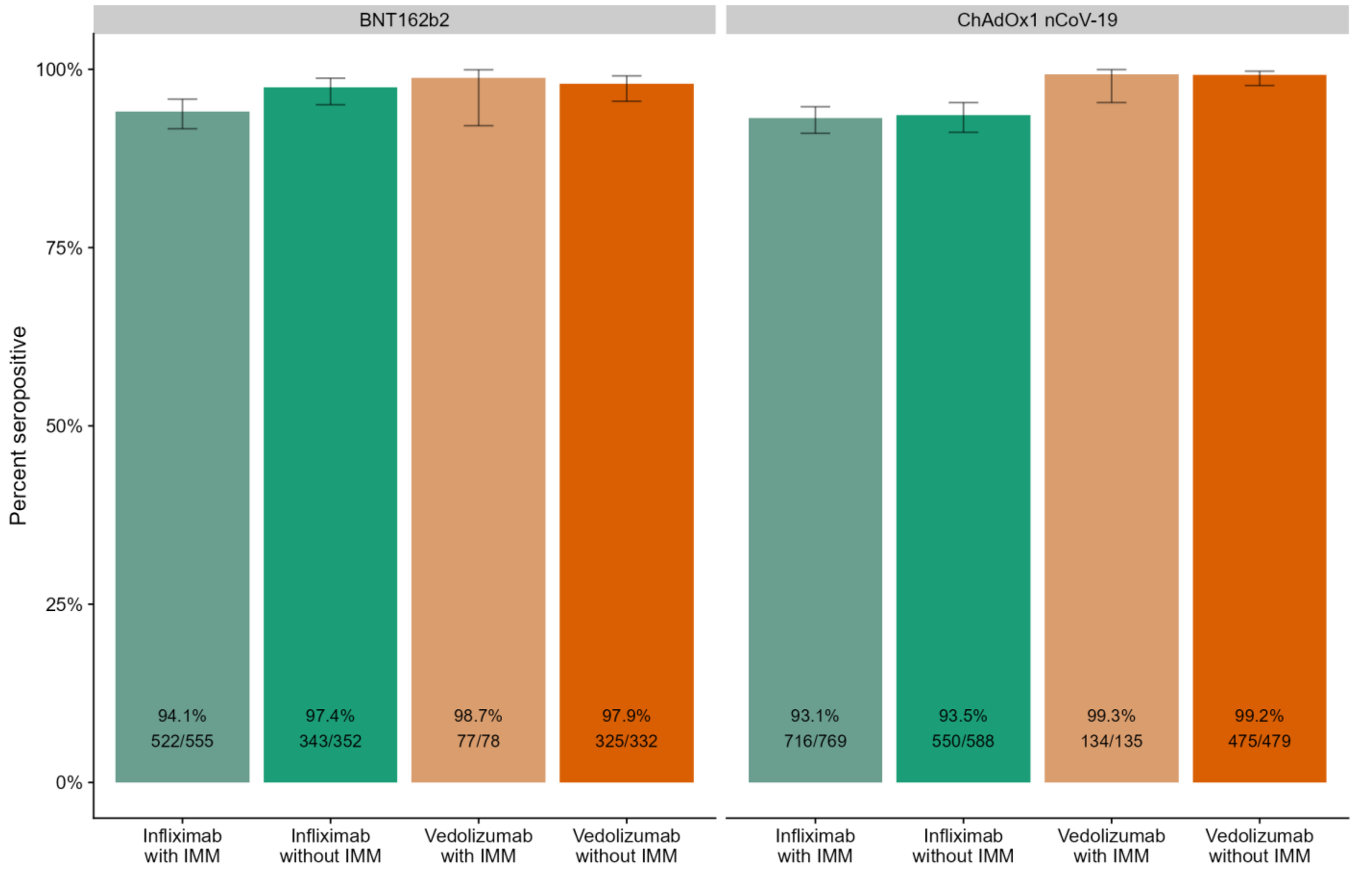

Supplementary Figure 2: Model estimates of the rate of anti-S RBD antibody decay over time, stratified by biologic therapy (infliximab vs vedolizumab), vaccine, and history of prior SARS-CoV-2 infection. λ represents the half-life calculated from the inverse of the gradient of a linear mixed effects model using log anti-S RBD antibody as the dependent variable and individual as a random effect.

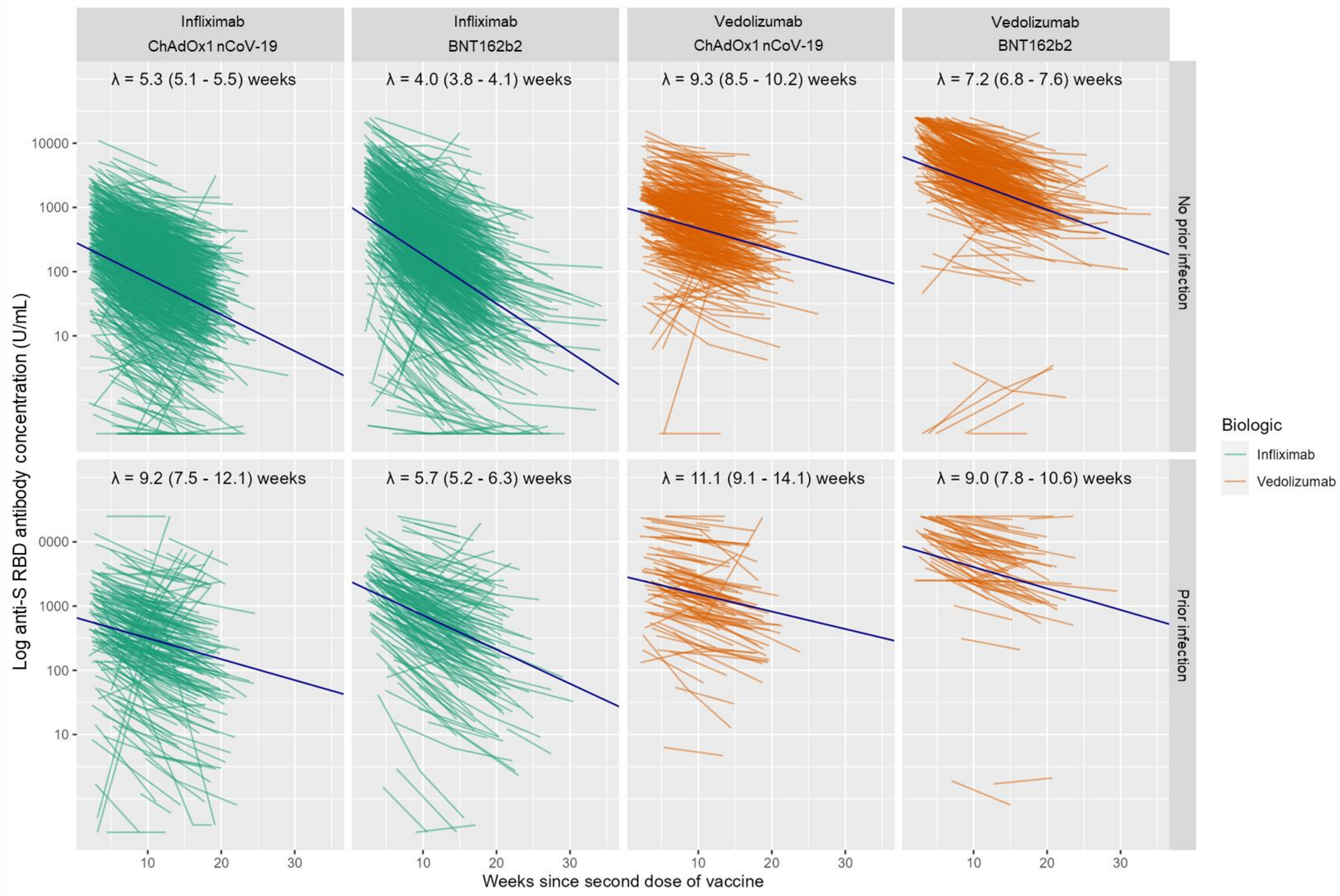

#### Supplementary Figure 3: Line graph showing the number of participants included at each time point in Figure 5 of the manuscript

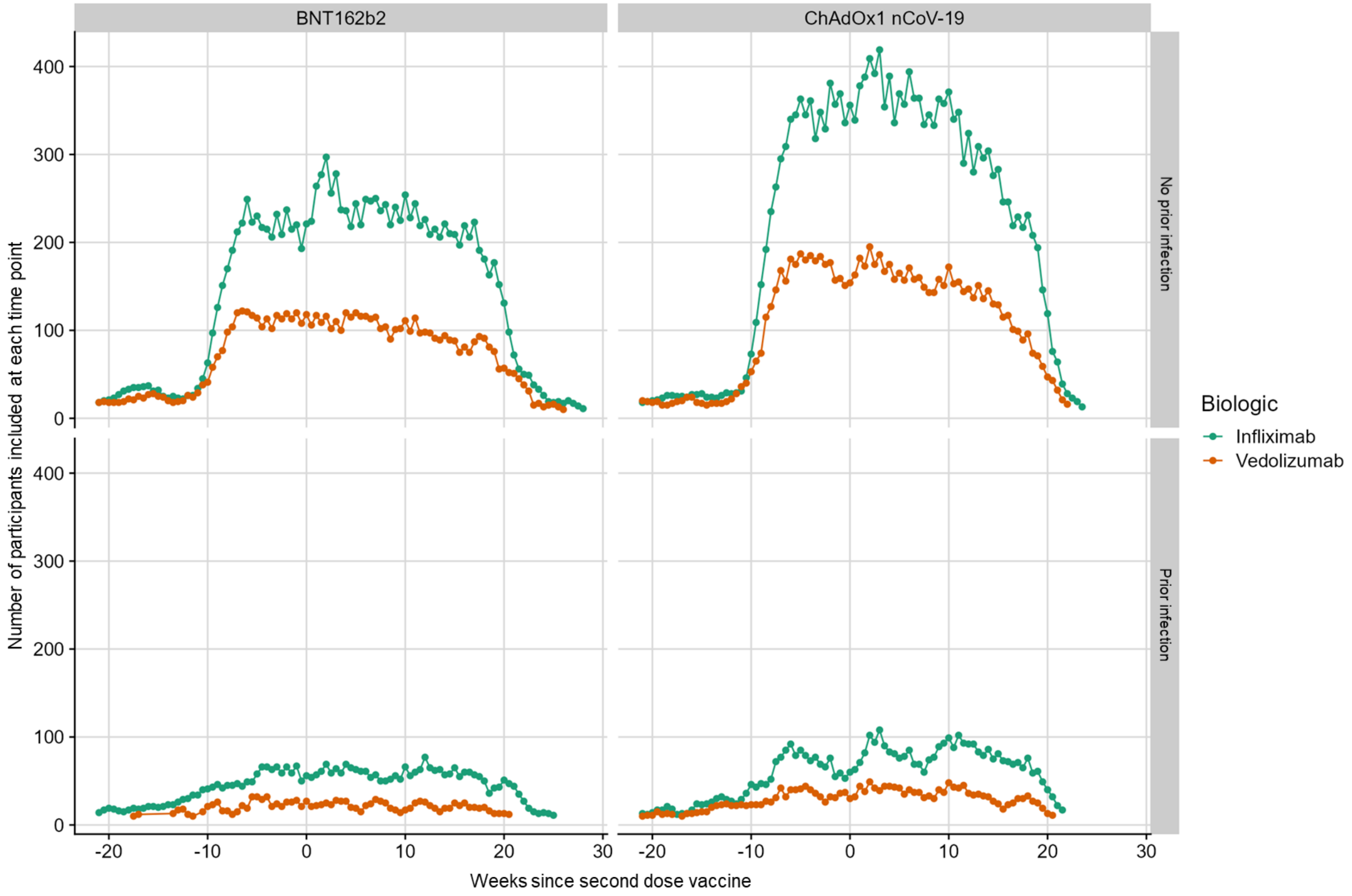

Supplementary Figure 4: Rolling geometric mean antibody concentration over time of participants on biologic therapy (infliximab vs vedolizumab) compared with 605 participants of the Virus Watch community cohort vaccine, timed from second dose SARS-CoV-2 vaccine (week 0), stratified by vaccine and history of prior SARS-CoV-2 infection.

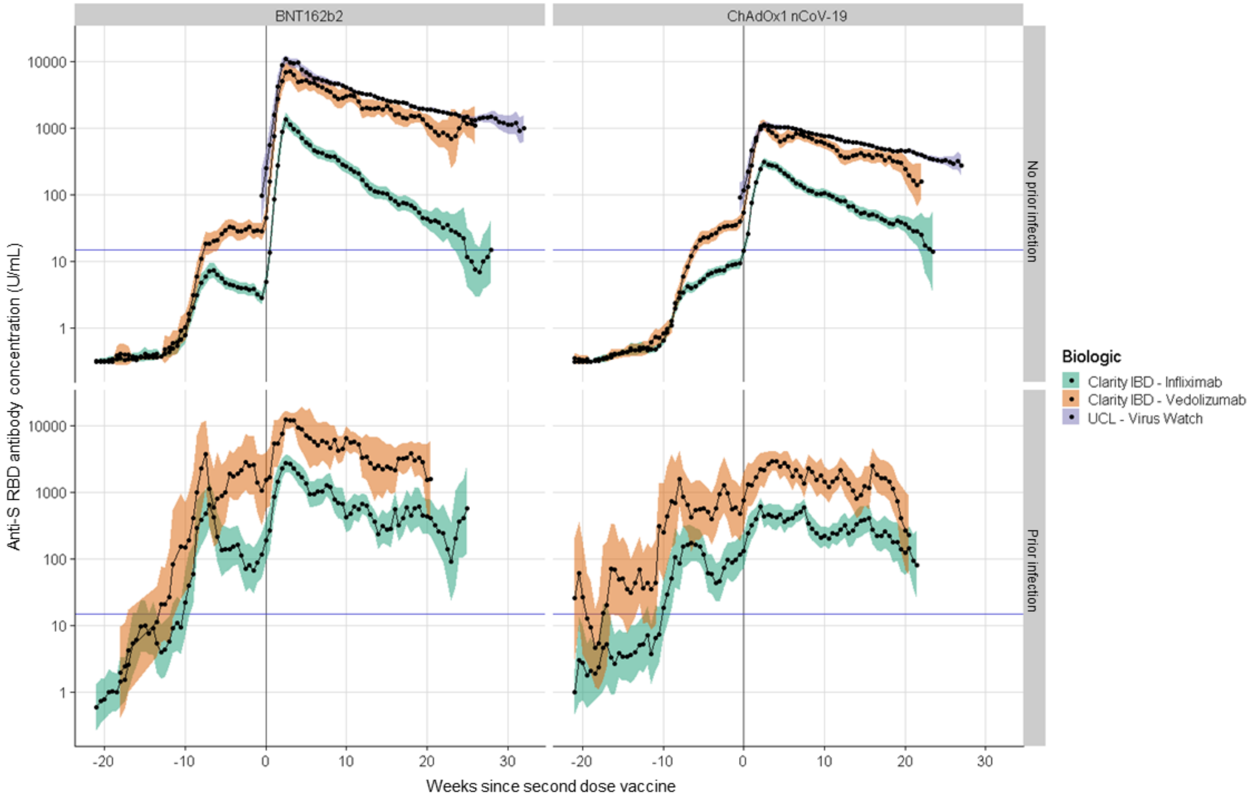

### Supplementary Figure 5: Kaplan-Meier graphs showing the time to the anti-S RBD antibody concentration falling below the seroconversion threshold of 15U/mL following the second dose of SARS-CoV-2 vaccine

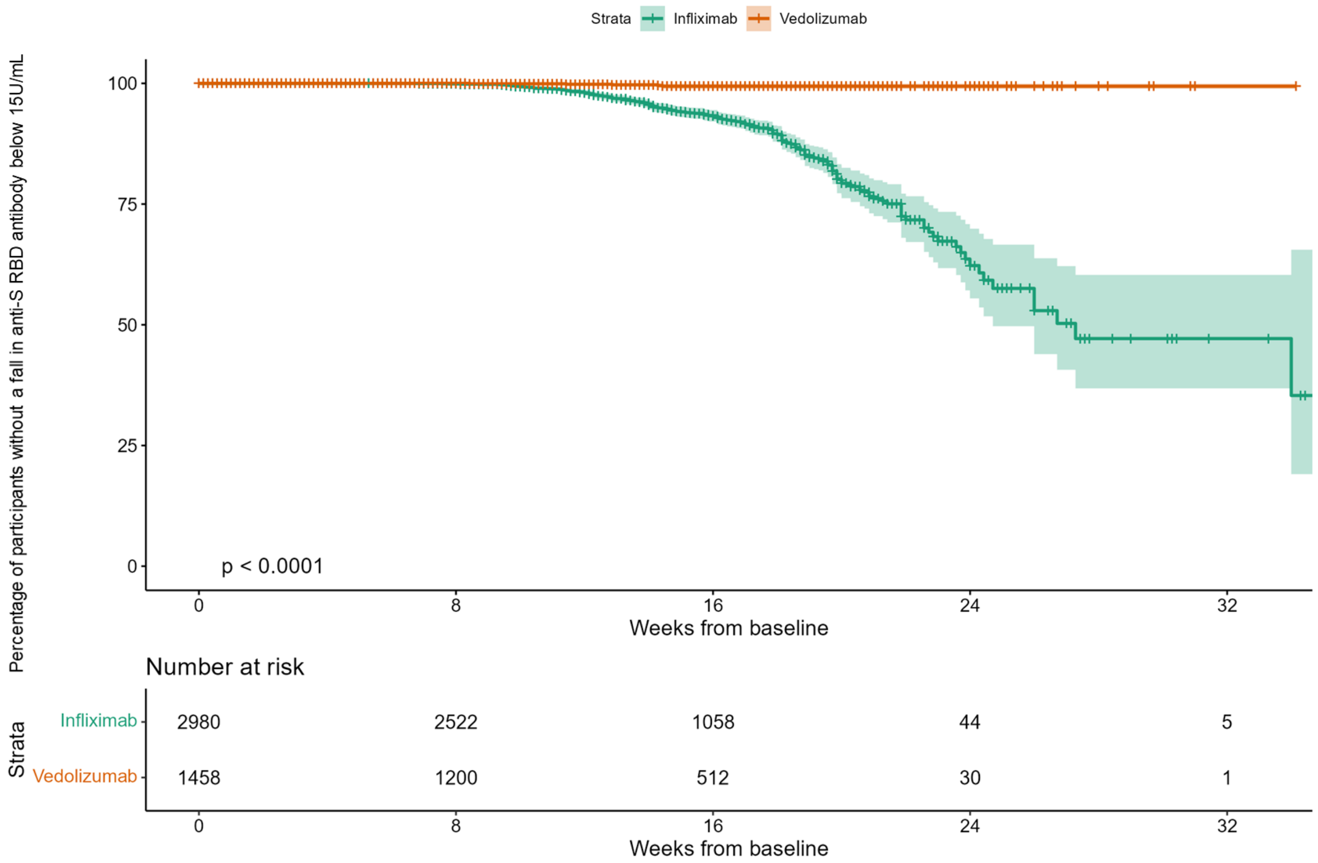

### Supplementary Figure 6: Cox proportional hazards model of variables associated with anti-S RBD antibody concentration falling below the seroconversion threshold of 15U/mL after the second dose of SARS-CoV-2 vaccine

| Variable | | N | Odds ratio | HR (95% CI) | p |
|---|---|---|---|---|---|
| **Biologic** | Vedolizumab | 1450 | | Reference | |
| | Infliximab | 2971 | | 21.70 (8.86, 53.13) | <0.0001 |
| **Immunomodulator** | | 2010 | | 1.06 (0.82, 1.38) | 0.66 |
| **Crohn's disease (vs UC or IBDU)** | | 2508 | | 1.23 (0.93, 1.63) | 0.15 |
| **Non-white ethnicity** | | 450 | | 0.40 (0.23, 0.70) | 0.0015 |
| **Current smoker** | | 418 | | 1.87 (1.32, 2.64) | 0.00044 |

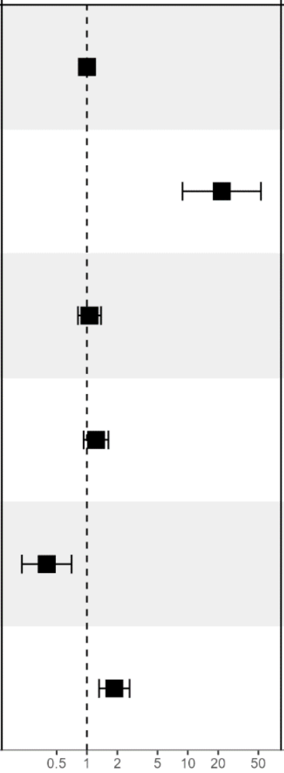

Supplementary Figure 7: Receiver operator characteristic curve of anti-N antibody results from participants two weeks following a PCR-confirmed infection. A threshold of 0.12 times the cut-off index provides 100% specificity for determining prior SARS-CoV-2 infection.

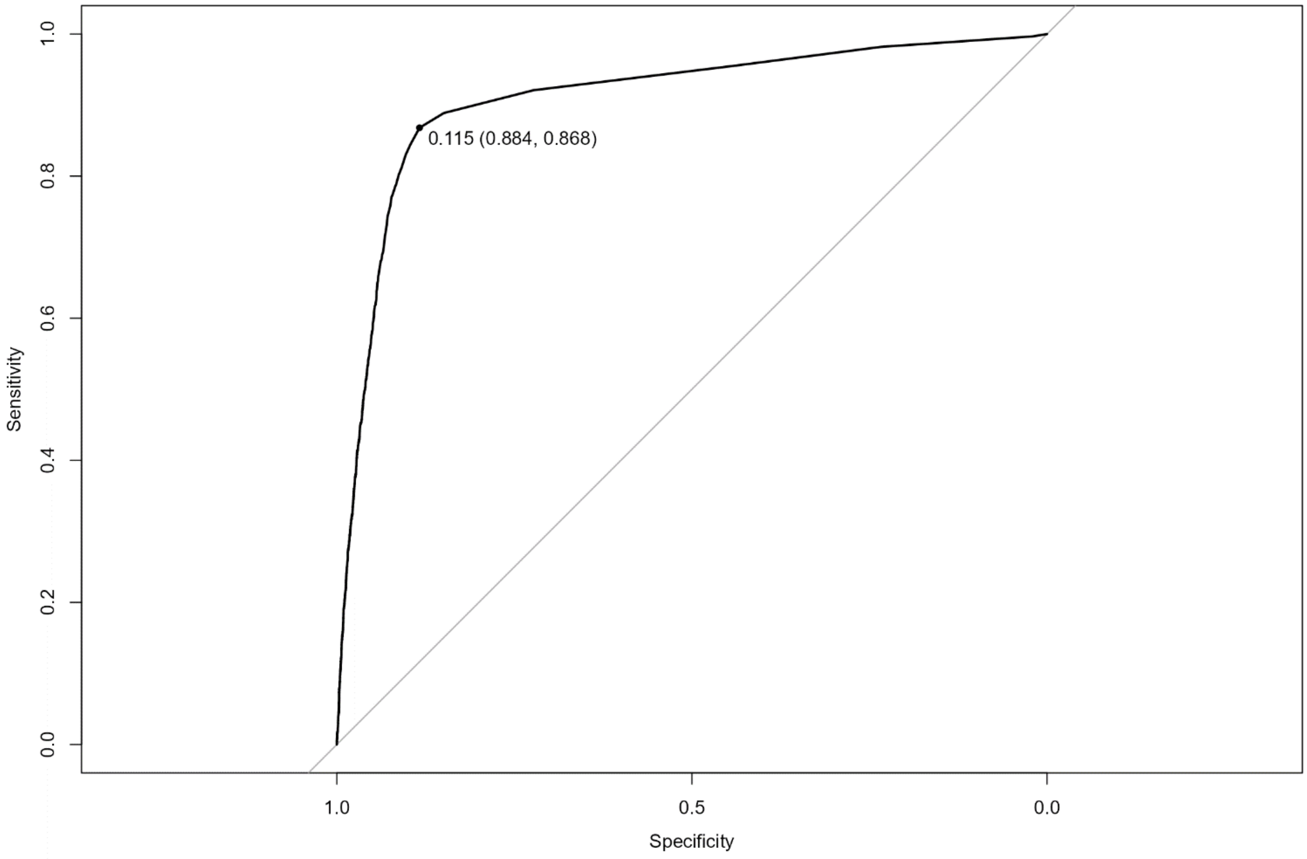

### Supplementary methods

In-house assay validation experiments on the Roche Elecsys Anti-SARS-CoV-2 spike (S) immunoassay were performed on 20 samples from healthy individuals who have been vaccinated. This demonstrated:

i) The intra-assay and inter-assay coefficient of variation were 1.3% and 5.6%, respectively

ii) Anti-SARS-CoV-2 (S) antibodies were stable in uncentrifuged blood and serum at ambient temperature for up to seven days permitting postal transport

iii) No effect was observed on recovery of anti-SARS-CoV-2 (S) antibodies following four freeze/thaw cycles

iv) No analytical interference was observed for the detection of anti-SARS-CoV-2 (S) with infliximab or vedolizumab up to 10,000 mg/L and 60,000 mg/L, respectively, or with anti-drug antibodies to infliximab or vedolizumab up to 400 AU/mL and 38 AU/mL, respectively (data not shown).
